# Estimating the genetic distance between subtypes of Major Depressive Disorder and their relationships with other traits using GDIS

**DOI:** 10.1101/2025.11.18.25340484

**Authors:** Anaïs B. Thijssen, Yuri Milaneschi, Meike Bartels, Brenda W.J.H. Penninx, Joëlle Pasman, Karin J.H. Verweij, Wouter J. Peyrot

**Author notes:** **Corresponding authors:** Anaïs B. Thijssen, Wouter J. Peyrot.

## Abstract

Investigating the genetic heterogeneity of psychiatric disorders, such as Major Depressive Disorder (MDD), is a key area of research, aimed at increasing etiological insights and ultimately improving clinical outcomes via personalized treatment. Previous research has used genome-wide association studies (GWAS) to investigate genetic heterogeneity by analyzing disorder subtypes versus controls (e.g. depression with comorbid anxiety versus controls, and depression without comorbid anxiety versus controls). However, the most important comparison for disentangling subtypes is typically neglected: the direct comparison subtype1-cases versus subtype2-cases (e.g. depression with comorbid anxiety versus depression without comorbid anxiety). We hypothesize that the direct comparison is being disregarded because of the lack of a generalizable distance metric (as observed or liability scale heritability are not readily interpretable), and because the data for this comparison may be harder to collect. Here, we introduce a novel method, Genetic DIstance of disorder Subtypes (GDIS), which benchmarks the genetic distance between subtype1-cases and subtype2-cases, by computing genetic distances (as the square root of the heritability on the 50/50 ascertainment scale) and geometrical angles (the inverse cosine of the genetic correlation). GDIS is readily applicable as it requires only subtype versus control summary statistics, and no individual-level data or GWAS of the direct subtype comparison. First, using data from the UK Biobank, we extensively validate the GDIS-geometrical representation to benchmark subtype1-cases versus subtype2-cases. Second, we applied GDIS to seven subtype-definitions of MDD to estimate the genetic distance between subtypes, informing their potential for future prediction and clinical stratification. Third, we extended and applied GDIS to investigate which subtypes were differentially correlated with each of 15 external traits; important information that cannot be distilled from the usual subtype vs control comparisons. Finally, GDIS aids intuition of the genetic interrelatedness of the subtypes through geometric visualizations. In conclusion, GDIS is a novel tool to benchmark direct subtype comparisons, yielding new insights into genetic heterogeneity of MDD.

## INTRODUCTION

The last decades have seen a tremendous increase in sample sizes for discovery of genetic variants associated with Major Depressive Disorder, resulting in new knowledge about the underlying genetic architecture and biology^1, 2^. Increases in sample size are most efficiently achieved by adopting a simple design of contrasting individuals with a diagnosis of MDD to healthy controls. However, a diagnosis of Major Depression can be made on the basis of many different constellations of symptoms^3^, making depression a far from uniform construct. This phenotypic heterogeneity complicates both fundamental research on the biological substrates of depression and likely contributes to the high variability of treatment success. To address this issue, research on the genetics of MDD has focussed on investigating subgroups of individuals that share key symptoms or clinical features. Analysing more homogenous samples can help with finding new, and more specific genetic risk loci, even at the cost of reducing sample sizes^4^. Moreover, it has the potential to help understanding the aetiology of different types of depression and aid in personalized medicine^5^.

Genetic heterogeneity is typically investigated by analyzing disorder subtypes (e.g. depression with/without comorbid anxiety), based on GWASs of each subtype versus controls. Typically, these two GWASs are then compared in terms of their heritability and genetic correlation with each other, and with other traits. Despite the great progress in this research^7, 8^, these types of analyses are unable to fully resolve the genetic differences between disorder subtypes, and are thus limited in their ability to acquire a complete understanding of disorder heterogeneity. Specifically, the direct genetic comparisons between subtype1-cases (e.g. MDD with comorbid anxiety) and subtype2-cases (e.g. MDD without comorbid anxiety) are generally disregarded, while these contain the most important information about the genetic distinctiveness of the subtype. We hypothesize that the direct comparison is being disregarded because of the lack of a generalizable distance metric, as the observed or liability scale heritability cannot directly be compared to each other due to scale differences. In addition, it might be more difficult to collect the needed individual-level data for this comparison.

Here, we introduce the Genetic DIstance of disorder Subtypes (GDIS) tool to benchmark and investigate direct subtype comparisons, and to intuitively visualize the heritability and genetic correlation estimates in geometrical representations. We apply GDIS to 7 previously defined subtypes of MDD^7^, contrast these subtypes to 15 external traits, and contrast these subtypes to each other.

## RESULTS

### Overview of methods

GDIS determines and benchmarks the genetic relationships between two subtypes of a disorder (subtype1-cases and subtype2-cases) and controls, aiding interpretation of their genetic differences and, optionally, differences in the relationship to an external trait. GDIS computes these relationships efficiently; to infer seven parameters including the genetic distances of subtype1-cases vs. controls, subtype2-cases vs. controls, subtype1-cases vs. subtype2-cases, and all-cases vs. controls, as well as the three genetic correlations between each comparison, GDIS requires only the two GWAS results of subtype1-cases vs. controls and of subtype2-cases vs. controls. The GDIS framework aids interpretation of the genetic correlations and heritabilities for a single subtype definition (e.g., MDD with/without comorbid anxiety), and also facilitates comparing across multiple subtype definitions (e.g., MDD with/without comorbid anxiety and MDD with/without recurrent episodes). GDIS furthermore provides visualizations of the geometric representation of the subtypes.

GDIS transforms SNP-heritabilities and genetic correlations to genetic distances and genetics angles, respectively (Figure 1, Figure S1). We define genetic distance (*GD*) and genetic angle (*GA*) as:

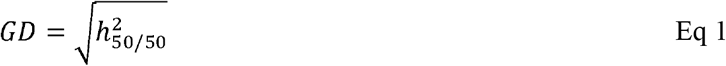

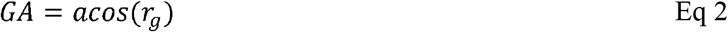

**Figure 1.**
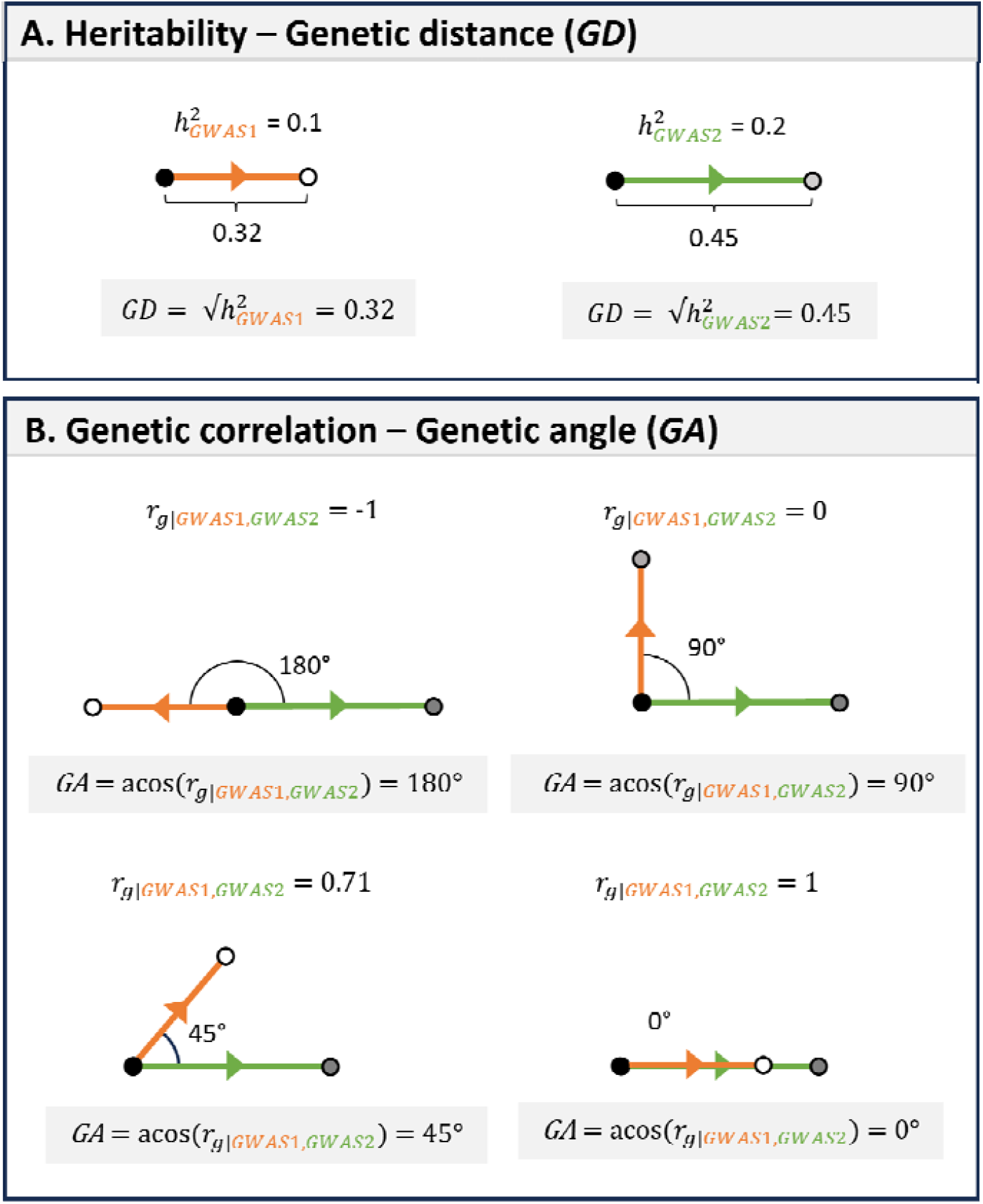
Transforming heritabilities to genetic distances and genetic correlations to genetic angles. Panel A. The square root of the heritability on the observed scale with 50/50 ascertainment ( ) provides a Euclidian distance measure between the two compared groups; the genetic distance follows the direction from the GWAS of the class coded as 0 to the class coded as 1 (the direction is relevant for the angle). **Panel B**. The genetic correlation between two GWASs corresponds to the inverse cosine of the angle formed by the line segments of these comparisons. Note that the inverse cosine gives the angle in radians, but we report the angle in degrees. GWAS1/GWAS2: GWAS comparing two classes (e.g., cases to controls, subtype vs. subtype or subtype vs. controls).

Specifically, we show that the square root of the heritability on the observed scale with 50/50 case-control ascertainment 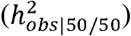 naturally corresponds to distance between the (sub)groups compared (see Methods; Figure 1a). The scale with 50/50 ascertainment provides a natural unified measure to evaluate the different subtype vs. control and subtype vs. subtype comparisons, because it does not rely on different population prevalences (as with the liability scale) or unbalanced ascertainment (as with the regular observed scale). For a comparison of the liability scale to the observed scale with 50/50 ascertainment based on ref.^9^, see Figure S2. Second, we show that the genetic correlation between the GWASs of two comparisons corresponds to the inverse cosine of the angle formed by the lines of these comparisons^10^. For example, a genetic correlation of 0 corresponds to an angle of 90 degrees (see Methods; Figure 1b).

GDIS uses nine steps to compute the geometric representation of genetic relations (Figure 2). First, LDSC^11, 12^ is applied to compute 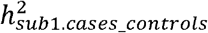 based on the GWAS results of subtype1-cases vs controls (*GWAS*_*sub1*.*cases_controls*_ ), 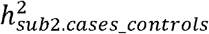 based on the GWAS subtype2-cases vs controls (*GWAS*_*sub2*.*cases_controls*_), and the genetic correlation *r*_*g*|*sub1*.*cases_controls,sub2*.*cases_controls*_ between these two comparisons. All heritabilities presented in this paper are on the observed scale with 50/50 ascertainment, which is attained by applying *N*_*effective*_ in LDSC. Second, the genetic distance of each subtype to the controls follows from the square roots of their heritabilities and defines two line segments (orange and green line in Figure 2). Third, the genetic angle formed between these line segments is determined by the inverse cosine of the genetic correlation (black dot in Figure 2). Fourth, the genetic distance between subtype1-cases and subtype2-cases (purple line in Figure 2) is computed with the law of cosines; note that squaring this genetic distance gives the heritability of this comparison. Fifth, the two remaining genetic angles of the triangle (at subtype1-cases and subtype2-cases; white and gray dots in Figure 2) are computed by again applying the law of cosines; note that the cosine of these angles will give the genetic correlation (i.e.,*r*_*g*|*sub1*.*cases_controls,sub1*.*cases_sub2*.*cases*_ and *r*_*g*|*sub2*.*cases_controls,sub1*.*cases_sub2*.*cases*_ ). Sixth, the group all-cases (i.e., the combined set of subtype1-cases and subtype2-cases; black square in Figure 2) lies on the line segment between subtype1-cases and subtype2-cases, where the distance between all-cases and subtype2-cases is inversely proportional to the proportion of subtype2-cases among all-cases (i.e., the larger the proportion of subtype2-cases, the closer all-cases is to subtype2-cases; Figure S1). Seventh, the genetic distance from all-cases to controls can be inferred using the law of cosines (gray line in Figure 2). Eighth, with all the distances known, the remaining angles (corresponding to *r*_*g*|*sub1*.*cases_controls,all*.*cases_controls*_ and *r*_*g*|*sub2*.*cases_controls,all*.*cases_controls*_) that are created with the line segment from all-cases to controls can be inferred from the law of cosines as well. Ninth, the population mean lies on the line segment between all-cases and controls, where the distance between population mean and all-cases is inversely proportional to the prevalence of all-cases (i.e., the higher the prevalence, the closer all-cases is to is to the population mean; asterisk in Figure 2). GDIS employs filters on the input to only compute the framework for subtypes that are genetically different based on *r*_*g*|*sub*.*cases_controls,sub2*.*cases_controls*_ being significantly different from 1 and/or significant different values of the heritabilities 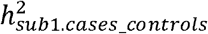 and 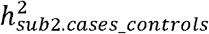. All GDIS-derived genetic distances and genetic correlations are validated with empirical analyses (see below).

**Figure 2.**
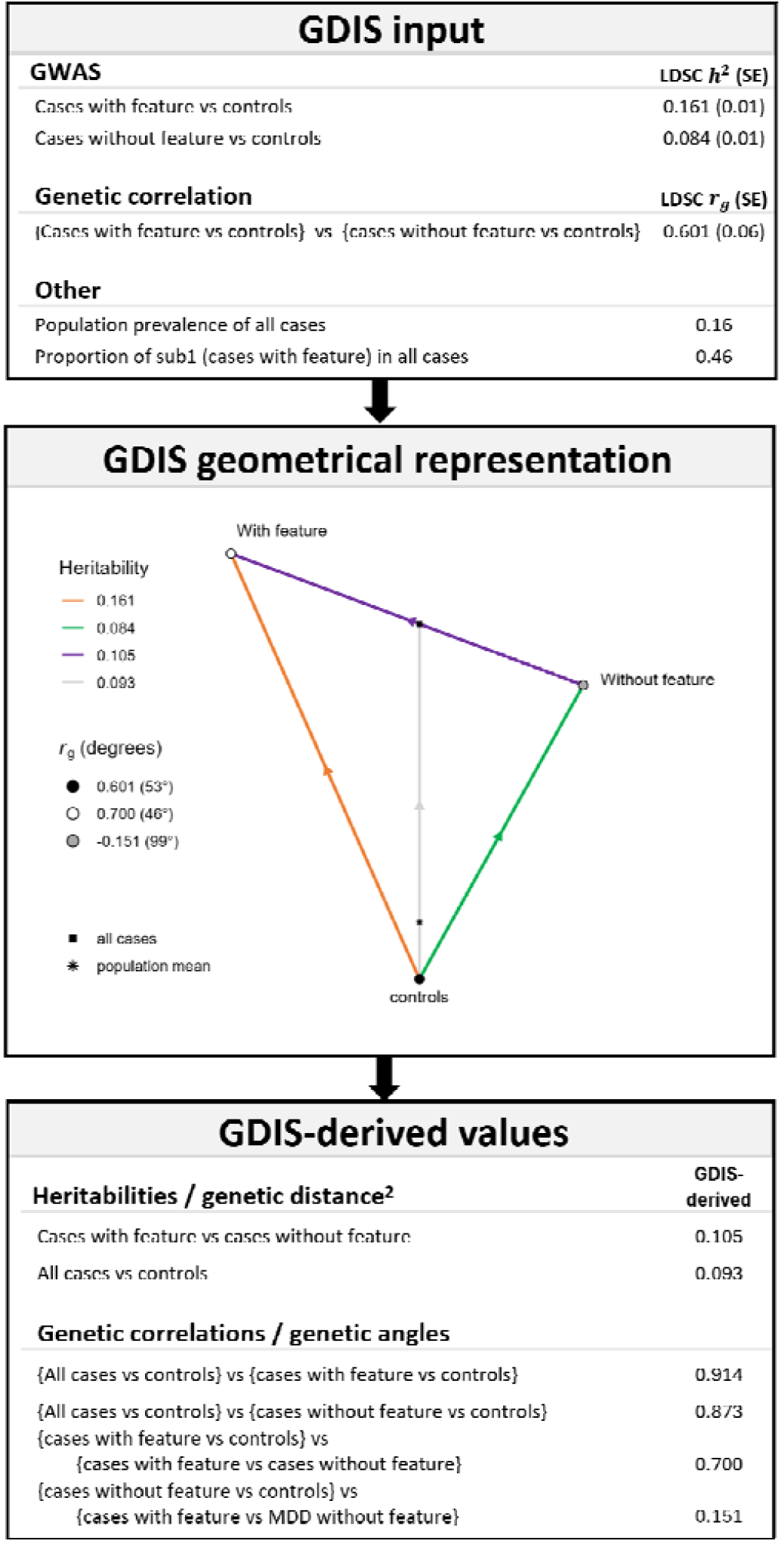
Overview of the GDIS workflow for a geometrical representation of a subtype. We report the GDIS-input, the GDIS geometrical representation of one subtype definition, and the GDIS-derived values based on geometrical computations. Heritabilities are on the 50:50 scale, the genetic distance (length of the line segments) corresponds to the square root of the heritability. With feature = subtype1-cases, without feature = subtype2-cases. The estimates in the example are for MDD with/without childhood trauma in Figure 3.

**Figure 3.**
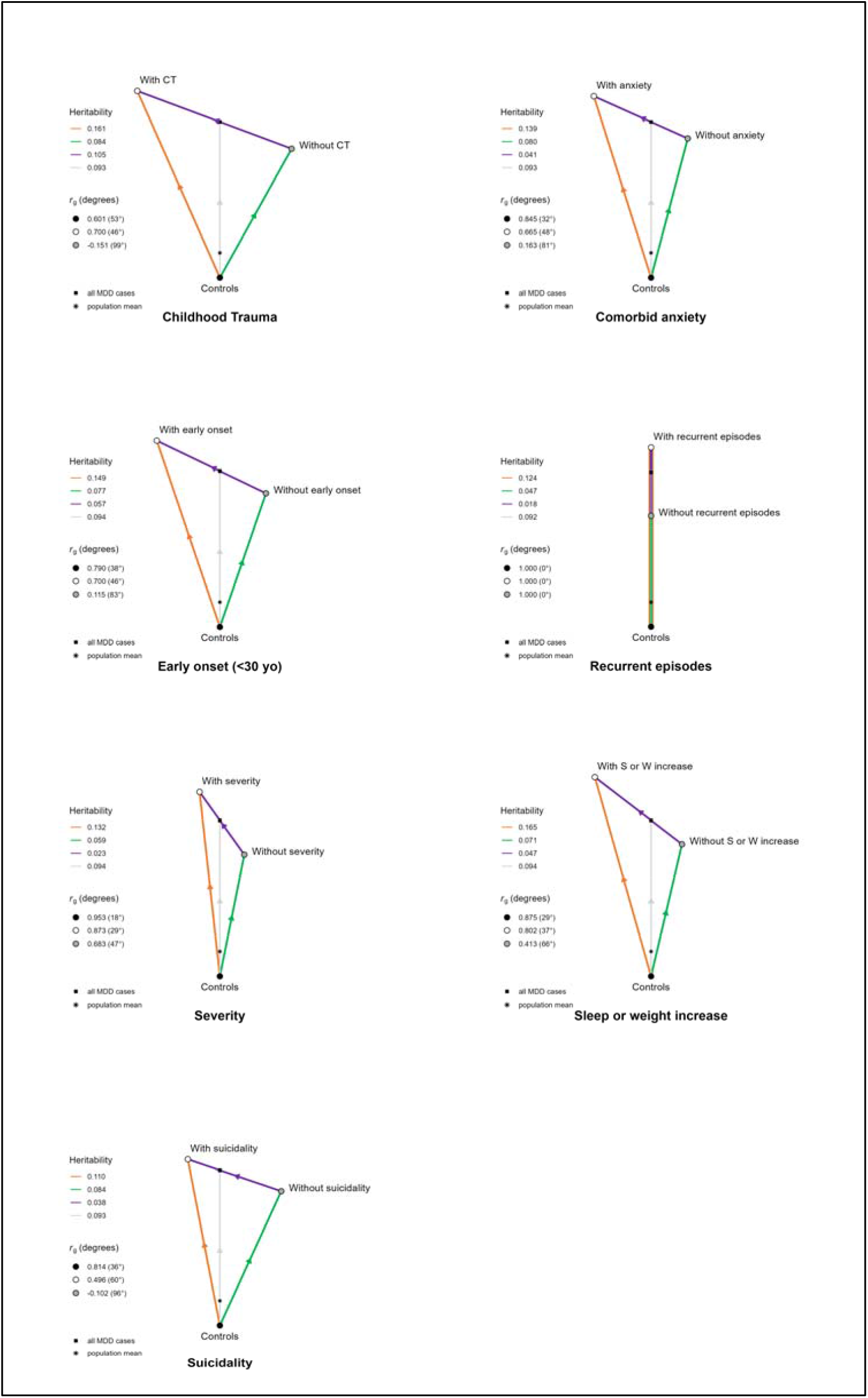
GDIS visualizations of seven major depression disorder subtypes. We present the GDIS visualization of the following seven subtypes of Major Depressive Disorder (MDD): with (subtype1-cases)/ without (subtype2-cases) childhood trauma, comorbid anxiety, early onset (< 30 yo), recurrent episodes, high severity, sleep or weight increase, suicidality. Numerical values are reported of input and GDIS-derived values are listed in Table 1.

GDIS builds on *F*_*ST,causal*_ (included in CC-GWAS^10^), a previous method for geometric representations of genetic distances (see Methods). However, *F*_*ST,causal*_ is a purely theoretical construct for independent causal SNPs, and its estimates are not intended as genome-wide metric nor has it been validated with empirical data. Furthermore, *F*_*ST,causal*_ does not extend into 3D geometric space nor does it include the relation with external traits.

We note that GDIS refers to the genetic distance as square root of the heritability, but there is an important conceptual difference between genetic distance and heritability: genetic distances have an interpretation between any two (potentially overlapping) groups, while heritability describes the variance explained by genetic effects in a phenotype in the population.

The geometric GDIS-representation of the subtypes can be extended to include an external trait (*ext*), resulting in a 3D geometrical representation. For this, GDIS additionally requires the GWAS of the external trait, which can be either a case-control trait (e.g. schizophrenia) or a continuous trait (e.g. BMI). First, 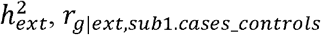, *r*_*g*|*ext,sub1*.*cases_controls*_ and *r*_*g*|*ext,sub2*.*cases_controls*_ are estimated with LDSC and converted to distances and angles respectively (see above, Figure 1, and Methods). While the angles have a clear interpretation for continuous traits, we note that the length of the line segment has no direct interpretation as genetic distance for continuous traits. The line segment of the external trait will intersect the subtype triangle at the population mean, resulting in a different placement of the controls of the external trait relative to the controls of the subtype triangle, reflecting that these groups of controls are not equal to each other. For both initial trait and external trait, the population mean lies on the respective case-control line segments, with its distance from the cases inversely proportional to the population prevalence. When the external trait is continuous, the population mean lies at the midpoint of the line representing the external trait. GDIS will derive estimates for *r*_*g*|*ext,all*.*cases_controls*_ and *r*_*g*|*ext,subl*.*cases_sub2*.*cases*_, which are validated in empirical analyses (see below).

GDIS can also be used to compare two subtype-definitions to each other in a 3D-geometric representation (e.g. subtype A: MDD with/without sleep or weight increase vs. subtype B: MDD with/without comorbid anxiety). The required input of this GDIS extension are the GWASs of subtype1-cases vs. controls and subtype2-cases vs. controls, for subtype-definition A and subtype-definition B (i.e. 4 GWASs in total). First, GDIS computes the representations for both single subtype definitions as above. Next, the genetic correlations between the subtypes are used to infer the relation between them, and are translated into corresponding genetic angles to form the geometrical representation. GDIS will derive estimates for *r*_*g*|*subA1*.*cases_subA2*.*cases,subB1*.*cases_subB2*.*cases*_, which are validated in empirical analyses (see below).

We applied GDIS to seven previously identified genetic subtype definitions of depression from the UK-Biobank, **with** (subtype1-cases) respectively **without** (subtype2-cases) the following features: childhood trauma, comorbid anxiety, early onset (first episode < age 30 years), recurrent episodes, high severity, sleep or weight increase, and suicidality (adapted from^7, 13^). Depression cases and controls were defined using the MHQ questionnaire^14^, yielding 30,431 MDD cases and 83,202 controls (Table S1). The subtype with the largest case-control unbalance was MDD with/without suicidality (*N*_*sub1*.*cases*_ = 19,998; *N*_*sub2*.*cases*_ = 10,433), while the most balanced was MDD with/without childhood trauma (*N*_*sub1*.*cases*_ = 14,146; *N*_*sub2*.*cases*_ = 16,285). For each subtype we ran two GWASs (with-feature vs. controls and without-feature vs. controls). To empirically validate the GDIS, we also performed the GWASs with-feature vs. without-feature, and the GWAS all-cases vs. controls. We considered GWASs of the following 15 external traits (Table S2), including binary traits: anxiety disorder (ANX)^13^, bipolar disorder (BIP)^15^, major depressive disorder (MDD)^2^, schizophrenia (SCZ)^16^, childhood trauma (CT)^17^; and continuous traits: body mass index (BMI)^18^, educational attainment (EA)^19^, Wellbeing^20^, Neuroticism^21^; and five Genomic-SEM cross-disorder factors: substance use dependence factor^22^, compulsive factor^22^, internalizing factor^22^, neurodevelopmental factor^22^, thought disorders factor^22^, and hierarchical factor^22^.

### Validating the GDIS-framework for subtype1-cases vs. subtype2-cases

We extensively validate the geometrical representation to benchmark subtype1-cases versus subtype2-cases. Applying GDIS to the depression subtypes, we show that GDIS-derived values match closely to LDSC-estimated values, with an average absolute difference of 0.002 (range 0 – 0.004) for the heritabilities (which are derived from the GDIS genetic distances) and 0.013 (range 0 – 0.077) for the genetic correlations (which are derived from the GDIS genetic angles; Table 1). The differences were slightly larger for the genetic correlations, with the larger differences corresponding to larger standard errors in the LDSC-estimates, while noting that all differences were within 0.59 standard error units of the LDSC estimate. We conclude that GDIS adequately benchmarks the subtype1-cases versus subtype2-cases genetic distance, based on input of only the GWAS results of subtype1-cases vs. controls and subtype2-cases vs. controls.

**Table 1.**
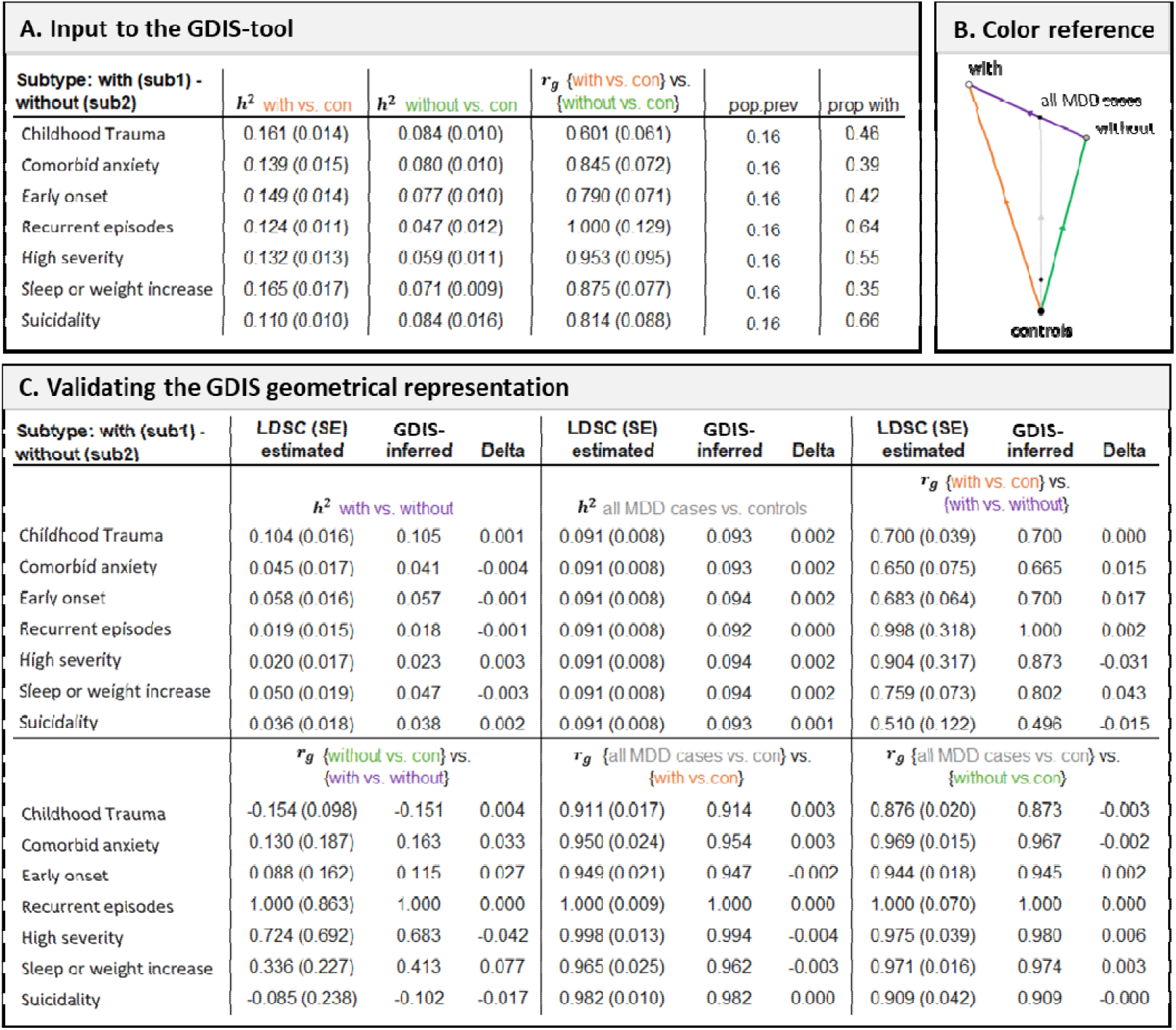
Validation of GDIS for the seven major depressive disorder subtypes. Panel A. Input for the visualization for GDIS. The heritabilities are on the observed scale with 50/50 ascertainment. Heritabilities and genetic correlations are estimated with LDSC. The population prevalence of major depressive disorder (MDD) is the same for each subtype. ‘Prop with’ refers to the proportion of MDD cases that have the ‘with’ subtype (e.g., MDD with childhood trauma). **Panel B**. Color reference for the comparisons presented in Panels A and C. **Panel C**. Comparison of GDIS-derived values vs. the LDSC-estimated values for the heritabilities of and, and the genetic correlations of these comparisons with the input GWASs.

### GDIS yields insights into several depression subtypes

As input, the heritability (i.e., genetic distance squared) of all-cases versus controls 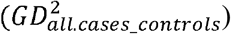 was 0.092 (se 0.008) as estimated with LDSC. After dividing the cases into subtypes, *GD*^2^ for the subtype with the feature (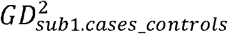 ranging from 0.110 (se 0.010) to 0.165 (se 0.017)) was consistently higher than that for the subtype without the feature (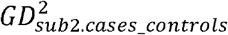 ranging from 0.047 (se 0.013) to 0.084 (0.010)), indicating that subtype without the MDD subtypes characterized by the presence of a given subtype-feature exhibit stronger genetic influences than cases in which that same feature is absent. The genetic correlations (i.e., the cosine of genetic angle) *r*_*g*|*subl*.*cases_controls,sub2*.*cases_controls*_ ranged from 0.601 (se 0.061) for childhood trauma to 1 (se 0.129) for recurrent episodes (as estimated with LDSC). GDIS computations revealed substantial differences in genetic distance of the direct comparison between the subtypes (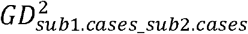 ranging from 0.018 for recurrent episodes to 0.105 for childhood trauma), demonstrating that some subtypes provide a more profound separation between MDD cases than others (Figure 3).

The genetic distance between subtype1-cases and subtype2-cases can result from (1) qualitative distinct genetic differences based on a *r*_*g*|*sub1*.*cases_controls,sub2*.*cases_controls*_ of less than 1 (e.g., mostly for the subtype-definition of suicidality); (2) a quantitative difference in magnitude of effects reflected in a difference between 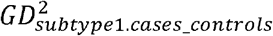 and 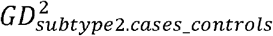 (e.g., subtype definition of high severity); or (3) a combination of both (e.g. subtype definition of early onset). The subtype definition based on childhood trauma showed the greatest genetic distance between subtype1-cases and subtype2-cases (*GD*^2^ = 0.105), which was even slightly larger than the genetic distance all-cases vs. controls (*GD*^2^ = 0.092). The GDIS-framework is thus able to intuitively condense a large number of estimates (including the 7×4=28 heritabilities and 7×5=35 genetic correlations) into seven subtype plots, which aids interpretation of this wealth of information (see Table 1, Figure 3).

Our main analyses were based on the UK Biobank MHQ definition^14^ without excluding related individuals to optimize sample size and power (see Methods). In secondary analyses, we explored the effect of using unrelated individuals and of using a broader depression definition. First, GDIS geometrical representations based on unrelated individuals were similar to those of related individuals (Figure S3). Second, we investigated the effect of using the broader depression definition as applied in the study by Nguyen et al.^7^, which furthermore uses mostly unscreened controls (see Methods). Here the GDIS geometrical representations were overall comparable, with slightly more pronounced differences for the subtypes of MDD with/without early onset and MDD with/without sleep or weight increase (Figure S4).

### GDIS compares subtypes to external traits

GDIS allows for the comparison of disorder subtypes to an external trait. We applied GDIS to investigate the genetic relation of the depression subtypes to 15 external traits (Table S2), including four dichotomous traits: anxiety disorder (ANX)^13^, bipolar disorder (BIP)^15^, major depressive disorder (MDD)^2^, schizophrenia (SCZ)^16^; five continuous traits: childhood trauma (CT)^17^, body mass index (BMI)^18^, educational attainment (EA)^19^, Wellbeing^20^, Neuroticism^21^; and six Genomic-SEM cross-disorder factors: substance use dependence factor^22^, compulsive factor^22^, internalizing factor^22^, neurodevelopmental factor^22^, thought disorders factor^22^, and hierarchical factor^22^.

The GDIS input consists of the GWAS of subtype1-cases, subtype2-cases and the additional GWAS for the external trait. GDIS only generates output after verifying that the genetic correlation estimates derived from these GWAS are compatible with each other (see Methods). The subtype recurrence with external traits did not pass this requirement, likely due to error terms in LDSC estimates (e.g., for anxiety disorder as external trait, *r*_*g*|*sub1*.*cases_controls,sub2*.*cases_controls*_ = 1 (se 0.129) cannot correspond with *r*_*g*|*ext,sub1*.*cases_controls*_ = 0.744 (se 0.192) and *r*_*g*|*ext,sub2*.*cases_controls*_ = 0.559 (se 0.223)). The external trait childhood trauma did not pass the checks for the subtype of childhood trauma and high severity. GDIS builds on the geometric subtype representation described above. To extend to the external trait, the genetic correlations *r*_*g*|*ext,sub1*.*cases_controls*_ and *r*_*g*|*ext,sub2*.*cases_controls*_ were computed using LDSC. Subsequently, GDIS derived estimates for *r*_*g*|*ext,sub1*.*cases_sub2*.*cases*_ using the rule of cosines and other geometrical computations (see Methods). We validated the GDIS-benchmarking framework by showing that the GDIS-derived estimates corresponded well to LDSC estimates, with an average absolute difference of 0.034 (range 0.0002 – 0.156; Figure 4, Table S3).

**Figure 4.**
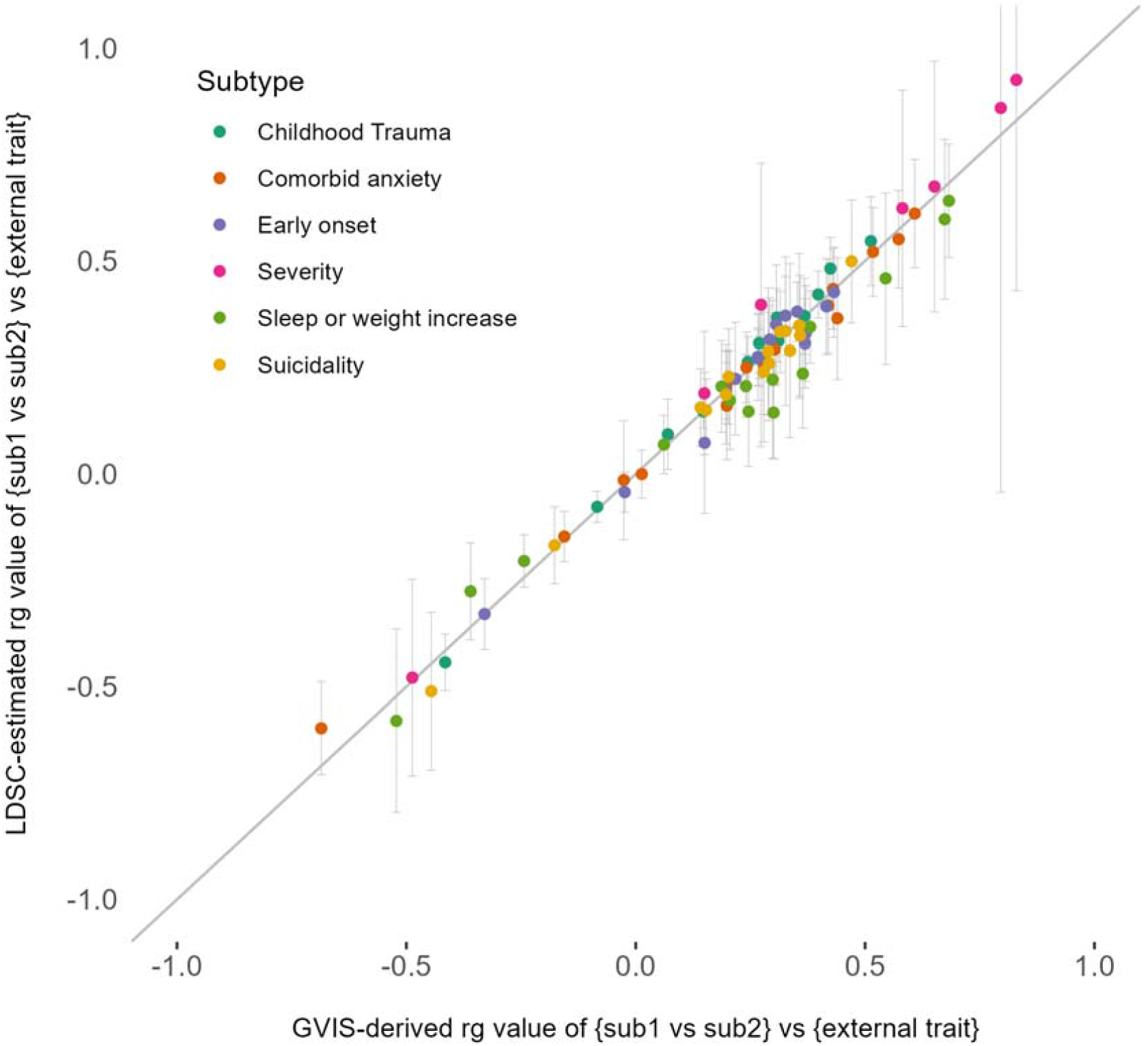
Validation of GDIS-derived values compared to LDSC-estimated values of the genetic correlations between external traits and subtype1-subtype2 comparisons. We report LDSC-estimated (x-axis) and GDIS-derived (y-axis) genetic correlation values for the external trait with the subtype1-cases and subtype2-cases comparisons ( *rg*|*ext,sub1*.*cases_sub2*.*cases* ). Different colors represent the six different subtypes that were related to 15 external traits (anxiety disorder^13^, bipolar disorder^15^, major depressive disorder^2^, schizophrenia^16^, childhood trauma^17^, body mass index^18^, educational attainment^19^, Wellbeing^20^, Neuroticism^21^, substance use dependence factor^22^, compulsive factor^22^, internalizing factor^22^, neurodevelopmental factor^22^, thought disorders factor^22^, and hierarchical factor^22^). Error bars represent ± the standard error of the LDSC estimate. The subtype of recurrence was not compatible for any of the 15 external traits and the external trait childhood trauma was not compatible for the subtype-definitions based on childhood trauma and high severity (due to incompatibility of LDSC input values, see main text). Numerical values for these results are reported in Table S4.

Several subtype-external trait pairs showed large differences between the genetic correlations *r*_*g*|*ext,sub1*.*cases_controls*_ and *r*_*g*|*ext,sub2*.*cases_controls*_ (Figure 5). For example, the correlation between the subtype MDD with weight increase vs. controls with BMI was 0.371 (se 0.032), while the subtype without weight increase vs. controls correlated 0.012 (se 0.036) with BMI. Most genetic correlations *r*_*g*|*ext,subl*.*cases_sub2*.*cases*_ were non-zero, ranging from - 0.878 for the comparison MDD with high severity vs. MDD without high severity to the external trait wellbeing, to 0.854 for the comparison MDD with high severity vs. MDD without high severity to the internalizing factor.

**Figure 5.**
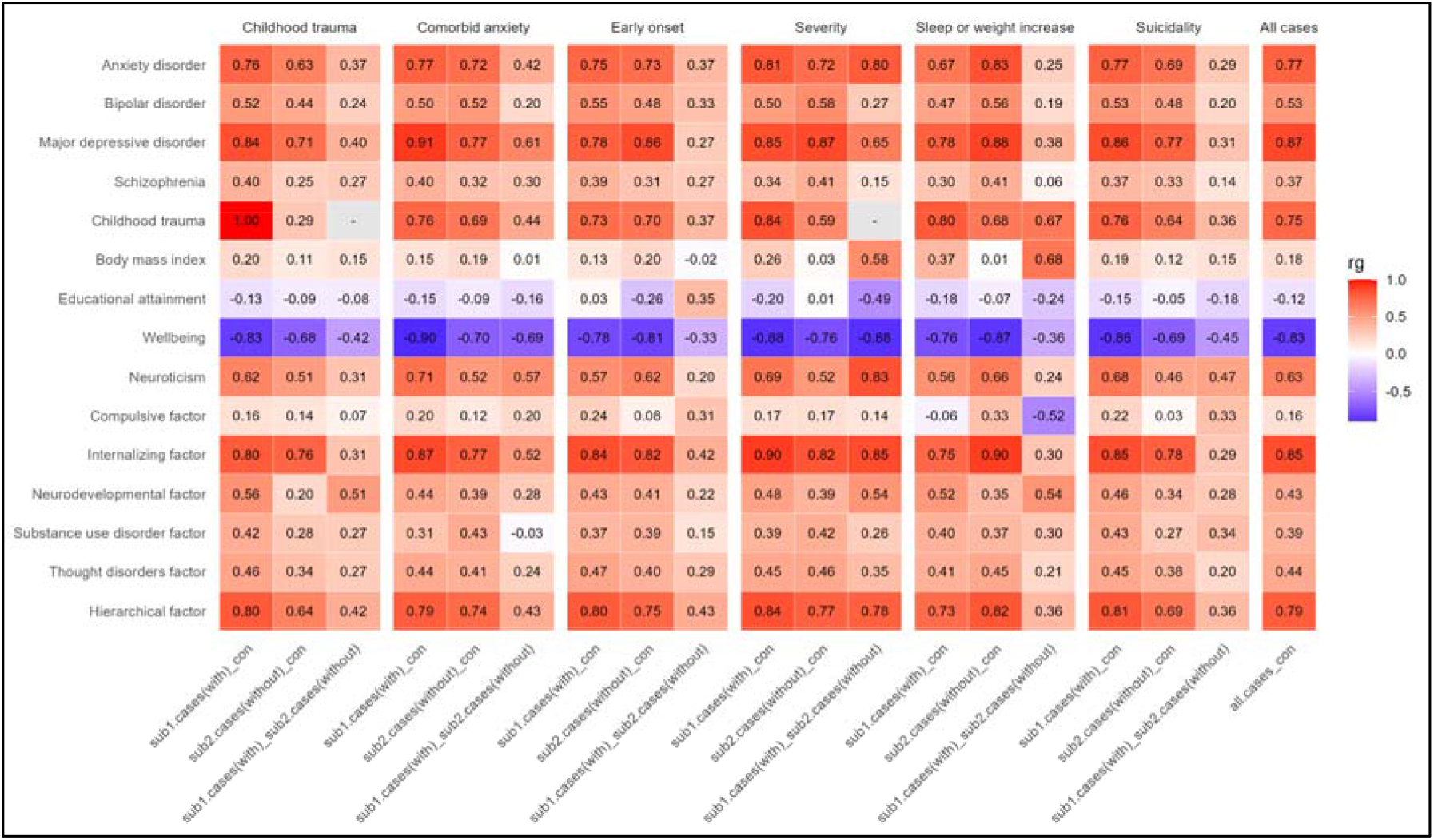
Genetic correlations of the subtypes with the external traits. We report the genetic correlation of the GWAS of 15 external trait with the following comparisons of six MDD subtypes (i) subtype1-cases versus controls, (ii) subtype2-cases versus controls, (iii) subtype1-cases versus subtype2-cases, and (iv) all-cases versus controls. The first two are based on LDSC and input to GDIS, the latter two are GDIS-derived. The subtype of recurrence was not compatible in geometrical space with external traits, the external trait childhood trauma was not compatible with the subtypes of childhood trauma and high severity (due to incompatibility of LDSC input values, see main text).

GDIS-results show that the direct subtype1-cases vs. subtype2-cases comparison provides important additional insight about the distinguishing potential of external traits on subtypes, that cannot be obtained by considering only the subtype1-cases vs. controls and subtype2-cases vs. controls comparison. For instance, for the subtype of high severity with the internalizing factor, the *r*_*g*|*ext,sub1*.*cases_controls*_ and *r*_*g*|*ext,sub2*.*cases_controls*_, do not reveal that the internalizing factor is a separating trait (i.e., a trait that is differentially associated with the subtypes), with values of 0.822 and 0.905, respectively. Still, *r*_*g*|*ext,sub1*.*cases_sub2*.*cases*_ is 0.854 (Figure 6A), indicating that the internalizing factor is differentially related to the subtypes. The 3D geometrical representations intuitively show that these seemingly contradictory findings are compatible, because they take into account how the subtypes interconnect. Specifically, the comparisons subtype1-cases vs. controls (orange line in Figure 6A), subtype2-cases vs. controls (green line), subtype1-cases vs. subtype2-cases (purple line) all form their own angle with the external trait (blue line), with the angle between subtype1-cases vs. subtype2-cases being driven by both the heritabilities of and the genetic correlation between subtypes1-cases vs. controls with subtype2-cases vs. controls. Figure 6B demonstrates the relation of the external trait BMI with the MDD subtype of with/without sleep or weight increase. BMI has a higher genetic correlation with the MDD with sleep or weight increase vs. controls (0.371) than with MDD without sleep or weight increase vs. controls (0.012), which is also reflected in the genetic correlation of the MDD with sleep or weight increase vs. MDD without sleep or weight increase vs. control with BMI (*r*_*g*_ = 0.683). This indicated that the subtype sleep or weight increase is closely genetically related to BMI, but is still distinct from BMI.

**Figure 6.**
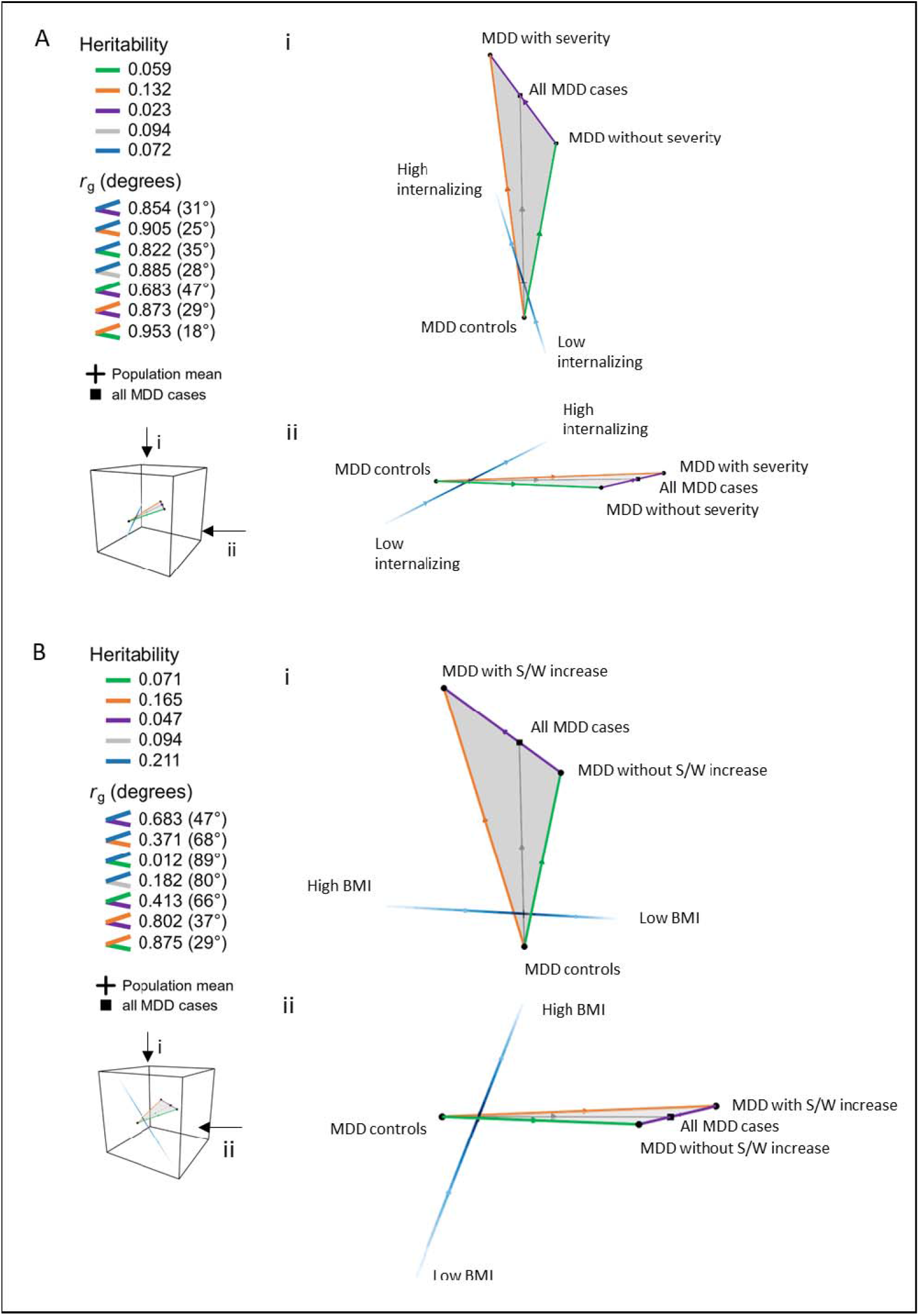
GDIS geometrical representations of subtypes with external traits. Panel A. GDIS geometrical representation of the major depressive disorder (MDD) subtype of high severity with the internalizing factor^22^ as external trait. **Panel B**. GDIS geometrical representation of the MDD subtype of sleep or weight increase with the external trait body mass index^18^. Heritabilities are on the 50:50 scale, the length of the line segments correspond to the square root of the heritability. For continuous traits, the length of the line segment has no direct interpretation as there are no clear endpoints. Visualizations exist in 3D geometrical space, as exemplified by the object in the cube. The top (i) and side (ii) views are taken to illustrate the relation of the subtype with the external trait. Interactive 3D visualizations can be accessed at: https://GDIS.shinyapps.io/GDIS/. All numerical values are reported in Table S3.

Geometrical GDIS representations of 88 subtypes vs. external traits are presented interactively at the GDIS shiny app: https://GDIS.shinyapps.io/GDIS/ (88 representations resulting from 7 subtypes x 15 external traits minus all the comparisons with the subtype recurrence and the two comparisons with the external trait childhood trauma that were not geometrically valid; see Methods).

### GDIS compares two subtype definitions to each other

We further extended GDIS to investigate how genetically distinct the subtype definitions were from each other. We excluded each subtype definition pair that was not compatible in geometric space resulting in 12 subtype pair comparisons (omitting all of the combinations with recurrence and most of the combinations with high severity; the two subtypes where *r*_*g*|*sub1*.*cases_controls,sub2*.*cases_controls*_ was not significantly different from 1; see Methods). To validate the GDIS-benchmarking framework, GDIS-derived values of *r*_*g*|*subA1*.*cases_subA2*.*cases,subB1*.*cases_subB2*.*cases*_ were compared to LDSC-estimated values (Figure 7, Table S4). The mean absolute difference was 0.05 (range 0.02-0.16), which was larger than the mean absolute difference for the comparisons with external traits; we note that this likely reflects the larger LDSC standard errors around these estimates due to smaller GWAS sample sizes, and that GDIS-derived values were all well within these standard error ranges. Results show that most subtypes were genetically divergent, as *r* _*g*|*subA1*.*cases_subA2*.*cases,subB1*.*cases_subB2*.*cases*_ values were not close to 1 (absolute range 0.072-0.732). For example, for the subtype definitions of comorbid anxiety and childhood trauma, *r*_*g*|*subA1*.*cases_subA2*.*cases,subB1*.*cases_subB2*.*cases*_ was 0.463, resulting in a cross-shaped geometric representation (Figure 8). GDIS geometric representations of the 11 other subtype-subtype comparisons can be found in Figure S5 and interactively in the GDIS shiny app. In secondary analyses, we compared the genetic correlations to phenotypic correlations; differences were found, but all phenotypical correlation estimates were within the 95% confidence interval of the genetic correlation estimates (Figure S6).

**Figure 7.**
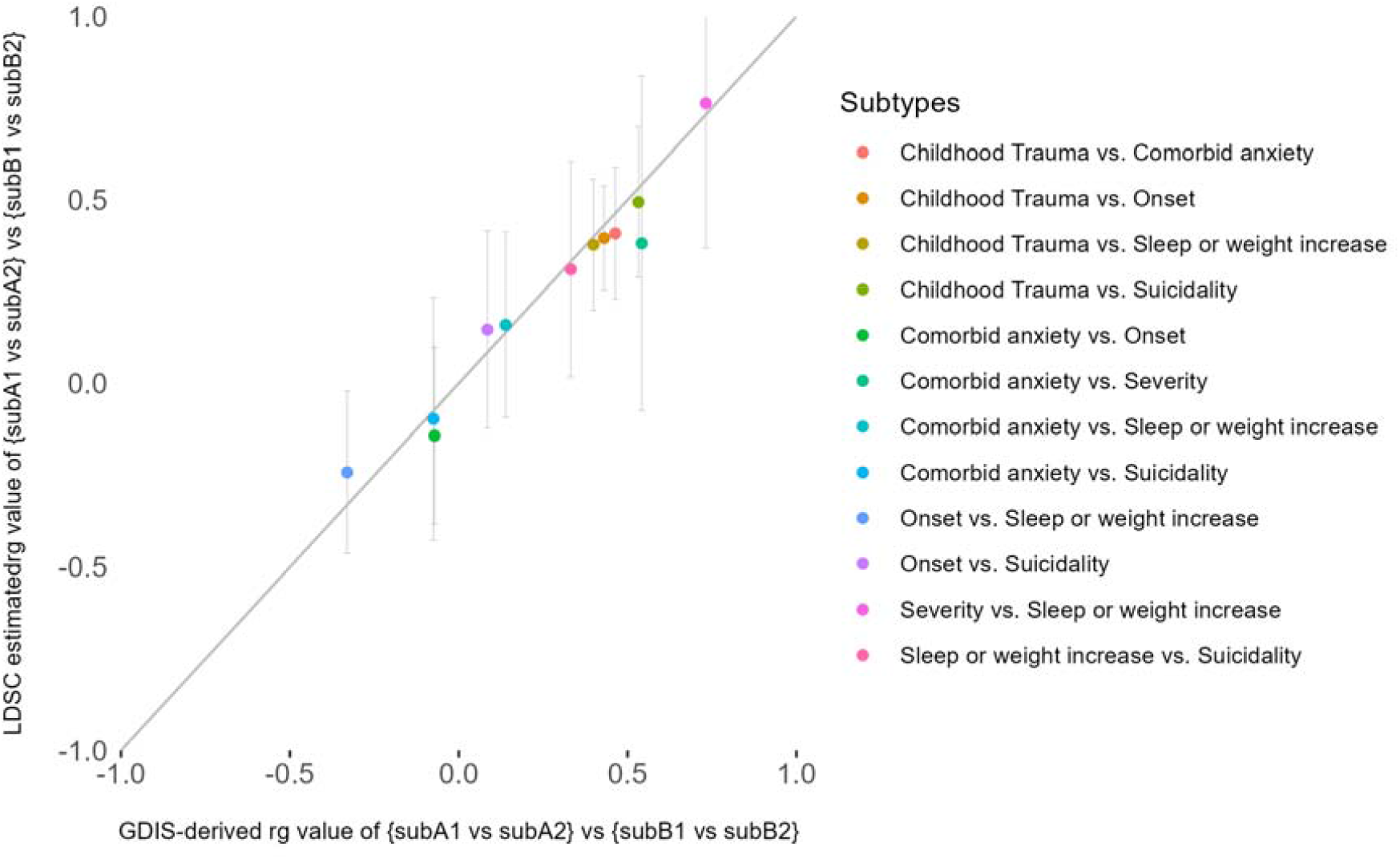
Validation of GDIS-derived values compared to LDSC-estimated values of the genetic correlation between the subtype1-subtype2 comparisons of two subtype-definitions. We report LDSC-estimated (x-axis) and GDIS-derived (y-axis) genetic correlation values for the comparisons of *GWAS* _*sub1*.*cases_sub2*.*cases*_ for subtype-definition A and subtype-definition B (e.g., *r* _*g*|*MDD*.*with*.*CT_MDD*.*without*.*CT,MDD*.*with*.*early*.*onset_MDD*.*without*.*early*.*onset*_ ). Error bars represent ± the standard error of the LDSC estimate. Different colors represent the different combinations of subtype-definitions. We exclude nine pairs of subtypes due to incompatibility for geometrical representation (see Methods). Numerical values are reported in Table S4.

**Figure 8.**
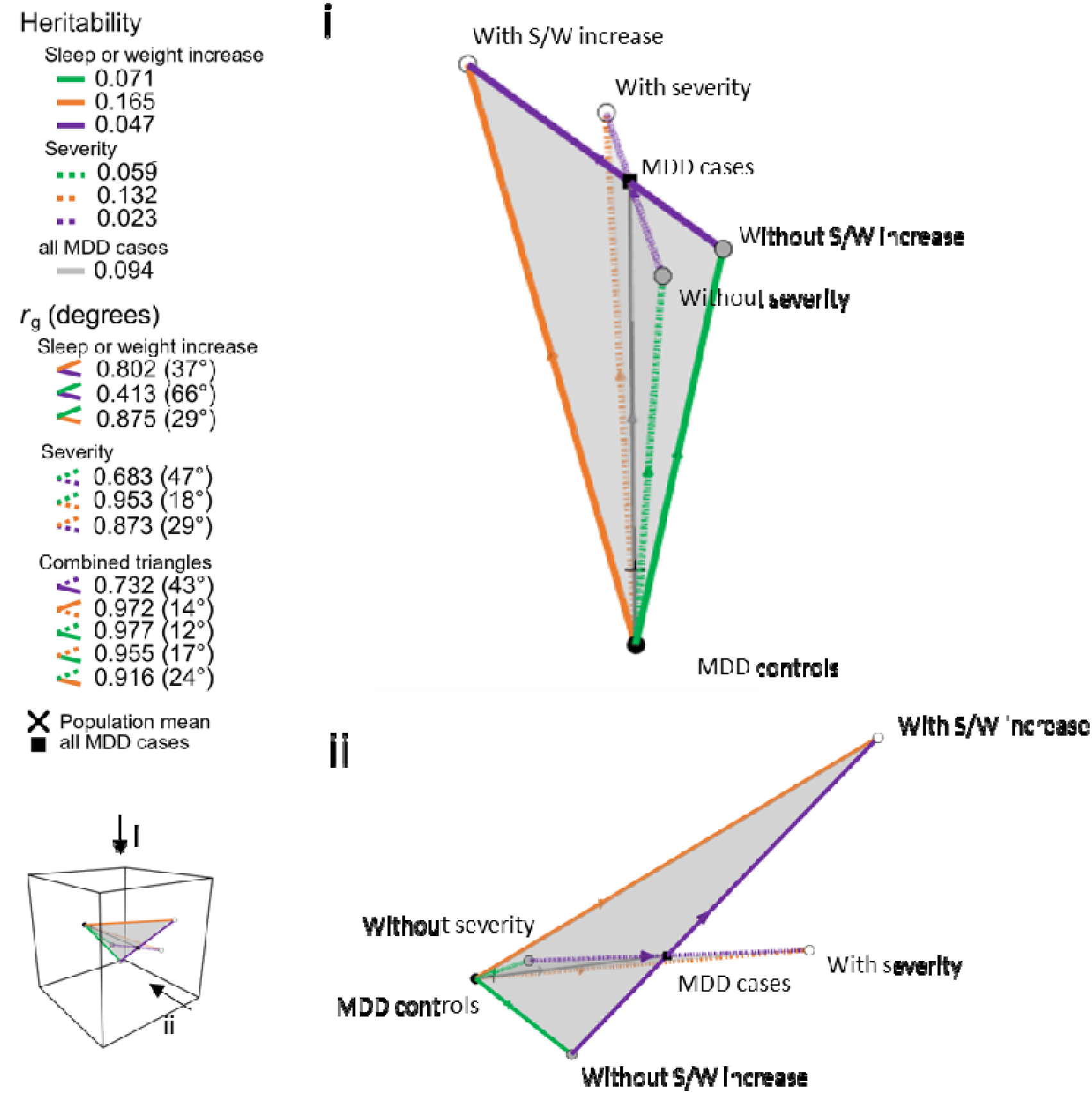
GDIS geometrical representation of the comparison of two subtype-definitions. We present the GDIS geometrical representation of the subtype-definition major depressive disorder (MDD) with/without high severity, and the subtype-definition MDD with/without high severity. The visualizations are in 3D geometrical space, as exemplified by the object in the cube. The top (i) and side (ii) views are taken to illustrate the relation between the subtype-definitions. Heritabilities are on the 50:50 scale, the length of the line segments correspond to the square root of the heritability. There is only one line segment representing the GWAS from MDD controls to all MDD cases, as these groups are the same in each subtype-definition. Interactive 3D visualizations can be accessed at: https://GDIS.shinyapps.io/GDIS/. All numerical values are reported in Table S4.

## DISCUSSION

We developed a new method, GDIS, to compute the genetic relationship between disorder subtypes, their relationship to external traits, and their interrelationship. GDIS benchmarks the genetic distance between subtype1-cases and subtype2-cases against heritabilities. Specifically, GDIS transforms heritabilities to genetic distances, and genetic correlations to genetic angles, resulting in geometric representations of the genetic architecture of the subtypes. We extensively validated all GDIS-derived estimates, which are based on the geometric representation of genetic relationships, as they are comparable to LDSC-based estimates. When comparing multiple subtypes, GDIS identifies which subtypes may offer the most potential for future clinical utility based on their genetic distinctiveness.

We applied GDIS to MDD subtypes, and make three main observations. First, the subtype1-cases vs subtype2-cases comparison, which contains information directly distinguishing subtypes, provides important added value over the subtype vs. controls comparisons alone (Figure 5 and 6). Specifically, the correlation of this direct comparison to external traits provides insight in how the subtypes are distinguishable—this cannot be uncovered from the respective subtype vs control comparisons, that largely capture the overall MDD signal. Second, by integrating the subtype1-cases vs. subtype2-cases relation with both subtype vs. control comparisons (Figure 3), GDIS shows that subtype1-cases vs. subtype2-cases genetic differences either arise from qualitative different genetic effects (i.e., *r*_*g*|*sub*.*cases_controls,sub2*.*cases_controls*_<1) or from quantitative different genetic effects (i.e., 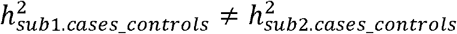). For example, the genetic difference between the subtypes with vs. without recurrent episodes reflects purely quantitative genetic effects, meaning that while the same risk alleles are involved in both subtypes, the magnitudes in the subtype with recurrent episodes are consistently larger across all SNPs. This finding is in line with theoretical predictions about the etiology of recurrent MDD episodes^23^. On the other hand, the most differentiating subtype is that of MDD cases with vs. without childhood trauma. Here, the genetic difference between the subtypes is mostly qualitative, with different genetic influences distinguishing the subtypes ( *r*_*g*|*sub1*.*cases_controls,sub2*.*cases_controls*_ = 0.60). While childhood trauma is an informative subtype due to its established clinical relevance^24-26^, we note that it is not a feature present only within MDD (as e.g., high severity of MDD), but also in controls, and caution that not all features present in both cases and controls (e.g. height) bear clinical relevance. Third, while GDIS-geometrical representations for single subtype-definitions might seem comparable (e.g. the clinically most severe subtype has the highest heritability across the subtypes), we show that the subtype definitions are genetically distinct (Figure 8). This finding parallels the low phenotypic correlations between the subtypes.

Multiple genetic methods exist to compare subtypes and disorders. First, a method often employed to evaluate genetic differences between subtypes involves estimating the heritabilities of the GWAS of subtype1-cases vs controls and subtype2-cases vs controls, as well as the genetic correlations between these two comparisons with external traits^7, 8^. GDIS derives and benchmarks the subtype1-cases vs subtype2-cases comparison, which adds crucial information on the genetic distinctiveness of the subtype. Second, GDIS builds on findings from *F*_*ST,causal*_ (included in CC-GWAS^10^), a previous method based on genetic geometric representations. However, *F*_*ST,causal*_ is a purely theoretical construct that is not empirically validated as GDIS. Furthermore, CC-GWAS is intended for association testing and not for genetic correlation estimation (specifically, the CC-GWAS case-case SNP-effect sizes have been optimized for power using two components (OLS and Exact), and it is unclear how these translate to case-case genetic distance and genetic correlation estimation, which will be biased towards the most powerful case-control GWAS in CC-GWAS). GDIS is much more versatile as it allows for the comparisons of disorder subtypes while extending into 3D geometric space to include the relation to external traits. In contrast to *F*_*ST,causal*_, GDIS infers the genetic distance based on all SNPs and not just independent causal SNPs. Third, a method applied to investigate genetic heterogeneity of a disorder or to compare different disorders is Genomic Structural Equation Modelling^22, 27, 28^. The goal of GSEM is different as it moves beyond the observed variables to create latent factors that summarize the coherence between symptoms or disorders. GSEM does not provide the direct geometric representation of heritabilities and genetic correlations like GDIS. Fourth, for subtype-features also present in controls, G x E interactions are a potential method to compare subtypes. For instance, research has addressed the question of whether the genetic risk for MDD interacts with childhood trauma^29^. GDIS focusses specifically on the difference between subtypes (in comparison to controls), whereas interaction analysis focuses on differences in direction and magnitude across environmental strata.

Findings from GDIS align with and expand on previous studies investigating genetic subtypes of MDD. First, a study investigating eight different depression subtype definitions relied on 16 subtype vs control GWASs^7^. They showed that three subtype definitions had distinct heritabilities and found differential genetic overlap of all subtypes with external traits. The study concluded that genetic heterogeneity exists within MDD and that genetic evidence shows that subtypes have partially distinct etiology. GDIS analyses extend this approach by using the subtype vs. subtype comparison and show that MDD subtypes are even more genetically differentiated than previously recognized. Furthermore, GDIS visualization were shown to be especially useful for intuitive comparison of multiple subgroups. Second, the subtype of typical versus atypical MDD has been of increasing interest in the literature^6, 30^. Research into the genetic underpinnings of this subtype have approximated the phenotype by focusing on hypersomnia and hyperphagia features. No significant heritability differences of the atypical vs control comparison and the typical vs control comparison have been found^8, 31^; however, as GDIS illuminates, the subtypes are genetically different when simultaneously taking the genetic correlation between these comparisons into account. This is supported by the polygenic score (PGS) analyses conducted in these previous studies, which showed distinct patterns of association with external traits^8, 31^. Third, age at onset of depression was recently investigated using a large Nordic dataset, contrasting early age at onset (< 25 years) with late onset (>50 years)^32^. As expected when comparing more extreme ends of the distribution, the subtypes were more genetically differentiated, with a genetic correlation of 0.58 compared to the genetic correlation of 0.79 that we obtained when splitting the age at onset as below or above 30 years. Consistent with our findings, the heritability of the early onset subtype vs. controls was higher than the late onset subtype vs. controls. The subtypes were compared directly in a subtype vs. subtype GWAS; however, the interconnectedness of the three GWASs was not fully resolved and difficult to intuitively comprehend as the heritabilities were expressed on different liability scales (rather than the observed 50/50 scale) and no genetic correlations were estimated of the subtype vs. subtype GWAS with the subtypes vs. controls GWASs or with external traits.

GDIS has several limitations. First, GDIS necessitates the use of heritabilities on the observed scale with 50/50 ascertainment and this scale is less widely used in the literature, limiting intuitive heritability interpretation. However, the benefit of this scale is that it provides a uniform distance measure for disorders with different population prevalences and for subtype vs. subtype comparisons. We provide Figure S2 to aid intuition of the relation between the 50/50 scale and the liability scale heritabilities. Second, GDIS does not compute standard errors, limiting the interpretation of the significance of results. However, the purpose of GVIS is not in significance testing, and we note that computing standard errors adds a disproportional layer of complexity due to the intrinsic challenges to compute standard errors of genetic correlations requiring block jackknife within LD-score regression^11^. Third, as has been exemplified by our application of GDIS to MDD, not all subtypes, subtype combinations, or relations between subtypes and external traits have a valid geometrical representation. However, this is either because there are no genetic differences between the subtypes, or because the LDSC-estimates are incompatible with each other. Furthermore, we note that empirical and derived values were in correspondence for the subtypes that do have valid geometrical representations. Fourth, although not a limitation of the method per se, we note that GDIS relies on which case and subtype definitions are used, which can remain slightly arbitrary and does have some impact on the geometrical representations (Figure S4). Fifth, although genetic space can be thought of as a >1,000,000-dimensional space (i.e. the number of independent SNPs), visual geometric representations are restricted to 3D space; hence, plotting more than one subtype and one external trait or more than two subtype-definitions is not possible. Sixth, the GDIS software is currently only able to compare two subgroups of a disorder that together form the complete set of cases. Therefore, GDIS is not suitable for subgroups that consist of three groups (e.g. low impairment, medium impairment, high impairment). A future research direction would be to extend GDIS to allow for comparison of three subgroups. Finally, this study made use of only individuals of European ancestry, thereby limiting the generalizability of findings to individuals of other ancestries.

In conclusion, GDIS provides a novel way of understanding the genetic architecture of disorder heterogeneity. GDIS is a freely available tool that can be used with any dichotomous subtype of a disorder. GDIS requires simple and minimal input data, which it uses to efficiently compute the geometrical representation, while automatically imposing checks and filters to ensure that only valid representations are produced. This makes the GDIS tool an invaluable tool in subtype research.

## METHODS

### The geometrical GDIS-representation of genetic distance and genetic correlation

GDIS transforms heritabilities to genetic distances, and genetic correlations to angles (Figure1). We define genetic distance (*GD*) and genetic angle (*GA*) as:

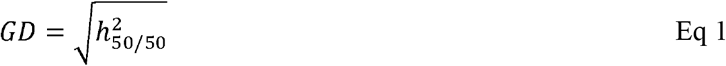

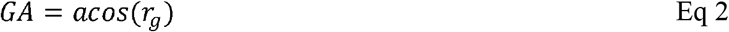

As demonstrated by the derivation of *F*_*ST,causal*_,^10^ the variance explained by a single causal SNP on the 50/50-ascertainment scale of a dichotomous phenotype 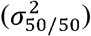 corresponds to the squared difference in standardized allele frequency, thus representing a Euclidian distance metric. Specifically, for genotype (*G*) and dichotomous phenotype (*D*) with 50% ascertainment,

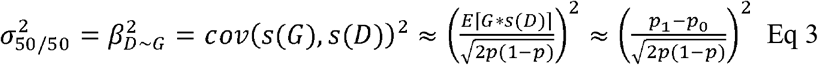

where *p* is the population allele frequency, *p*_1_ the frequency in cases, *p*_0_ the frequency in controls, and *s*(*G*) and *s*(*D*) are the standardization to a mean of 0 and variance of 1 for *D* and *G* respectively. Previous work derived this correspondence for independent causal SNPs^10^; here we show via extensive empirical analyses that this extends to a genome-wide correspondence of the square root of heritability as distance metric (Table 1, Figure 4, Figure 7). Thus, the GWAS of a dichotomous phenotype *D* can be represented as a line segment with length 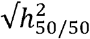 in Euclidian space.

When considering two GWASs of two dichotomous phenotypes (*D*_*A*_ and *D*_*B*_), they form two line-segments in Euclidian space. These two line-segments form an angle 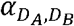:

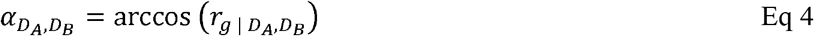

This equation has previously been derived for independent causal SNPs^10^; here we show via extensive empirical analyses that this extends to a genome-wide correspondence of genetic correlation estimated with LDSC^11^ (Table 1, Figure 4, Figure 7).

GDIS follows the same rationale as the genetic distance measure *F*_*sT,causal*_: the analytically derived *F*_*ST*_ at causal independent SNPs for cross-disorder case/control comparisons^10^. However, GDIS differs in the following key aspects. First, GDIS infers the genetic distance based on all SNPs and not just independent causal SNPs. Second, GDIS can be applied on disorder subtypes and not only for cross-disorder comparisons. Third, GDIS-estimates of genetic distances and angles are validated with empirical results of heritabilities and genetic correlation, while *F*_*ST,causal*_ is purely an analytical construct used to increase power of cross-disorder case vs. case GWAS.

Thus transforming heritabilities to genetic distances and genetic correlations to angles (Figure 1) allows for geometrical computations based on heritabilities and genetic correlations (see below).

### GDIS method for one subtype-definition

GDIS uses nine steps to compute the geometric representation of the genetic architecture of a subtype.

#### Step 1: Estimate heritabilities and genetic correlation

GWAS results of subtype1-cases vs controls (*GWAS*_*sub1*.*cases_controls*_) and subtype2-cases vs controls (*GWAS*_*sub2*.*cases_controls*_) are used to compute 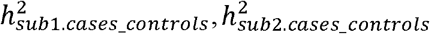 and *r*_*g*|*sub1*.*cases_controls,sub2*.*cases_controls*_ with LDSC (Bulik-Sullivan 2015a, 2015b). The heritabilities are expressed on the observed scale with 50/50 ascertainment, which is computed by using *N*_*effective*_ in LDSC. For unrelated individuals, *N*_*effective*_ is computed by 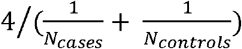 (refs.^11, 12, 33, 34^). For related individuals, *N*_*effective*_ for each variant is computed by 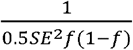, where is the allele frequency of the genetic variant and SE is the standard error of the effect of the variant in the linear mixed model from fastGWA^35, 36^. When the LDSC-estimated genetic correlation value exceeds 1, it will be capped at 1 for the geometrical computations.

#### Step 2: Compute genetic distances of subtype1.cases_controls and subtype2.cases_controls

Taking the square root of the heritabilities obtained from step 1 gives the genetic distance from subtype1-cases to controls (orange line) and subtype2-cases to controls (green line), i.e. the length of the line segments in the GDIS-representation.

#### Step 3: Compute the angle between subtype1.cases_controls and subtype2.cases_controls

The inverse cosine of *r*_*g*|*sub1*.*cases_controls,sub2*.*cases_controls*_ gives the genetic angle formed by the respective line segments at the controls in radians (represented by the black dot). We note that we report angles in degrees.

#### Step 4: Compute the sub1.cases_sub2.cases genetic distance

Using the length of the line segments from *subtype1*.*cases_controls* and *subtype2*.*cases_controls* and their angle, the length of the line segment *subtype2*.*cases_controls* (purple line) is computed with the law of cosines: 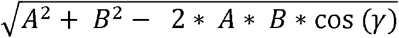 where A is the length of *subtype1*.*cases_controls*, B is the length of *subtype2*.*cases_controls* and *γ* is the angle between these lines. Squaring the resulting line length of *subtype1*.*cases_subtype2*.*cases* gives its GDIS-derived heritability (which is empirically compared to LDSC estimates; see below and Table 1).

#### Step 5: Compute angles with sub1.cases_sub2.cases line segment

The angle formed between the line segment of *subtype1*.*cases_controls* (orange line) and *subtype1*.*cases_subtype2*.*cases* (purple line) can be computed using the law of cosines: acos((*C*^2^ + *A*^2^ − *B*^2^) / (2*AC*))where C is the length of the line segment for *subtype1*.*cases_subtype2*.*cases*, A is the length of the line segment for *subtype1*.*cases_controls* and B is the length of the line segment for *subtype2*.*cases_controls*. The angle between the line segments of *subtype2*.*cases_controls* (green line) and *subtype1*.*cases_subtype2*.*cases*(purple line) can be computed similarly. Taking the cosine of the angle will give the genetic correlation.

#### Step 6: Compute all.cases_sub1.cases and all.cases_sub2.cases line segments

The all-cases group is the union of subtype1-cases and subtype2-cases. Note that for the *all*.*cases_controls* line segment to be equal to the overall disorder-controls line segment, the subtype1-cases and subtype2-cases together need to cover the complete set of cases. The point corresponding to the all-cases group all-cases lies on the line segment of subtype1-cases and subtype2-cases. The proportion of subtype1-cases to subtype2-cases determines where along the line the all-cases point falls. Specifically, the proportion of subtype1-cases to all-cases times the length of *subtype1*.*cases_subtype2*.*cases* determines the distance from all-cases to subtype2-cases: 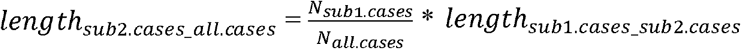 . For example, a larger proportion of subtype1-cases will result in all-cases being closer to the subtype1-cases point. The length of subtype1.cases to all-cases is the length of *subtype1*.*cases_subtype2*.*cases* minus the length of *subtype2*.*cases_all*.*cases*.

#### Step 7: Compute all.cases_controls genetic distance

To get the length of the line segment from all-cases to controls (gray line), the law of cosines can be used: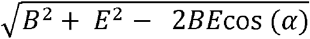, where B is the length of the line segment from subtype2-cases to controls (obtained in step 2), E is the length of the line segment from all-cases to subtype2-cases (obtained in step 6) and α is the angle between these lines in radians (obtained in step 5). Squaring this distance will give the heritability.

#### Step 8: Compute angles with all.cases_controls line segment

The angle formed between the line segment *all*.*cases_controls* and *subtype1*.*cases_controls* can be computed with the law of cosines: *acos* ((*F*^2^ + *A*^2^ − *D*^2^)/(2 ∗ *A* ∗ *B* )), where F is the length of *all*.*cases_controls*, A is the length of *subtype1*.*cases_controls* and D is the length of *all*.*cases_subtype1*.*cases*. The angle between the line segment *all*.*cases_controls* and *subtype2*.*cases_controls* can be computed similarly. The resulting angles are converted to genetic correlations by taking the cosine.

#### Step 9: Compute the population mean

The population mean lies on the line segment from all-cases to controls, where the distance from controls to the population mean is related to the population prevalence of the disorder (analogue to step 6). This distance is calculated by multiplying the length from *all*.*cases* to controls with the population prevalence.

#### Supplementary validation analyses

The *GWAS*_*sub1*.*cases_sub2*.*cases*_ will often not be available, and is not necessary as it can be computed from GDIS. However, when *GWAS*_*sub1*.*cases_sub2*.*cases*_ is available, supplementary validation analyses can be performed. The *GWAS*_*all*.*cases_controls*_ can also be used for validation, which is specifically relevant when it is uncertain if the union of sub1.cases and sub2.cases form to full set of cases (see step 6 above). The GDIS-software has the option to compare its derived values based on these GWASs to the LDSC-estimated values. Specifically, for 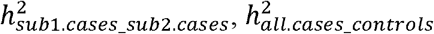, *r*_*g*|*sub1*.*cases_controls,sub1*.*cases_sub2*.*cases*_, *r*_*g*|*sub2*.*cases_controls,sub1*.*cases_sub2*.*cases*_, *r*_*g*|*all*.*cases_controls,sub1*.*cases_controls*_, and *r*_*g*|*all*.*cases_controls,sub2*.*cases_controls*_ GDIS determines what the difference between the GDIS-derived and the LDSC-estimated values are, and how the magnitude of this difference relates to the magnitude of the standard error of the. We present these validation steps for our analyses in Table 1.

#### Input filtering: check whether the subtypes are genetically distinct

Before creating the geometrical representation of a subtype, GDIS checks the input data to make sure that only heritable, genetically different subtypes are plotted. First, GDIS checks whether *GWAS*_*sub1*.*cases_controls*_ and *GWAS*_*sub1*.*cases_controls*_ are significantly different from 0. Next, GDIS checks whether there are indications that the subtypes are genetically distinct by determining (1) whether the 95% confidence interval of *r*_*g*|*subl*.*cases_controls,sub2*.*cases_controls*_ contains 1, and (2) whether the heritabilities of *GWAS*_*sub1*.*cases_controls*_ and *GWAS*_*sub1*.*cases_controls*_ are within each other’s 95% confidence intervals. If the genetic correlation is not significantly different from 1 and the heritabilities do not differ significantly, this suggests that the subtypes are not significantly different and no GDIS geometrical representations will be computed.

### GDIS method for one subtype-definition with an external trait

The geometric GDIS-representation of a subtype can be extended to include an external trait (*ext*). For this, GDIS additionally requires the GWAS of the external trait. To create the 3D-geometrical visualization, GDIS computes the subtype representation as before, and then follows these steps:

#### Step 1: Estimate genetic distances and genetic correlations

The heritability and genetic correlations 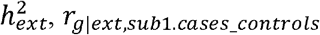, *r*_*g*|*ext,sub1*.*cases_controls*_ and *r*_*g*|*ext,sub2*.*cases_control*_ are estimated using LDSC. If the external trait is a dichotomous trait, 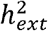 needs to be on the observed scale with 50/50 ascertainment, which can be computed as described in step 1 above. The external trait can also be a continuous trait, for which the heritability is expressed on its natural scale. For continuous traits, only the GDIS angles are of interest; the transformation of heritability to genetic distance is less informative because it does not connect defined groups like cases and controls. Next, the genetic correlation of the external trait with *GWAS*_*sub1*.*cases_controls*_ and *GWAS*_*sub2*.*cases_controls*_ is calculated using LDSC.

#### Step 2: Compute the ext.cases_ext.controls or external trait genetic distance

The line length from the external cases to external controls is obtained by taking the square root of the heritability from step 1. Note that the external controls are not the same as the controls used in the subtype triangle, which is reflected by the two points not being in the same geometrical space. When the external trait is continuous, the line segment will not connect cases to controls, and the ends are not well-defined, which is visually represented by a fading line.

#### Step 3: Compute the angles between the line segment of ext.cases_ext.controls with the line segments of subtype1.cases_controls and subtype2.cases_controls

The angle formed between the line segment *ext*.*cases_ext*.*controls* with *subtype*1.*cases_controls* and *subtype*1.*cases_subtype2*.*cases* can be computed by taking the inverse cosine of the genetic correlations calculated in step 1.

#### Step 4: Compute the population mean point for the external trait

The line segment of the external trait will intersect the subtype triangle at the population mean. To calculate where on the external trait line segment the population mean lies, the population prevalence is used as described above (in step 9 of the section ‘*GDIS method for one subtype-definition’*). For continuous traits, the population mean is represented by the middle of the line segment.

### Supplementary validation analyses

If the *GWAS*_*sub1*.*cases_sub2*.*cases*_ and/or *GWAS*_*all*.*cases_controls*_ are available, GDIS has the option to compare its derived values based on the GWASs to the LDSC-estimated values, also for the external trait. Specifically, for *r*_*g*|*all*.*cases_controls,ext*_, and *r*_*g*|*sub1*.*cases_sub2*.*cases,ext*_ it is determined what the difference between the GDIS-derived and the LDSC-estimated values are, and how the magnitude of this difference relates to the magnitude of the standard error of the LDSC estimate. We present these validation steps for our analyses in Figure 4 and Table S3.

### Input filtering: check compatibility for geometrical representation

Due to inherent variance of LDSC estimates, it may occur that the subtype and external trait are not compatible for geometric representation. Specifically, in some instances *r*_*g*|*sub1*.*cases_con,sub2*.*cases_con*_ can be very close to 1 (e.g., 0.99) while the correlation with the external trait ( *r*_*g*|*sub1*.*cases_con,ext*_ and *r*_*g*|*sub2*.*cases_con,ext*_) differ considerably (e.g., 0.5 vs 0.3). GDIS filters for such inconsistencies. First, GDIS computes a helper triangle and determines whether this is a valid triangle. The helper triangle is the base of a triangular pyramid which has the controls as apex with three sides going down from the controls to the base which are all 1. The three face angles at the controls correspond to *r*_*g*|*sub1*.*cases_con,ext*_, *r*_*g*|*sub2*.*cases_con,ext*_, and *r*_*g*|*sub1*.*cases_con,sub2*.*cases_con*_. For the base triangle to be a valid triangle, the sum of its smallest two lines has to be longer than the length of the longest line. If this is not the case, the base is not a valid triangle, indicating that the three angles *r*_*g*|*sub1*.*cases_con,ext*_, *r*_*g*|*sub2*.*cases_con,ext*_, and *r*_*g*|*sub1*.*cases_con,sub2*.*cases_con*_ are not compatible with each other, in which case GDIS will not plot the subtype with the external trait.

### GDIS method to compare two subtype definitions

GDIS can compare two subtype definitions (definition A and definition B) to each other by simultaneously computing two single subtype triangles in the same geometrical space. The required input is the same as the input for the single subtype triangles. GDIS only computes this geometric representation if the two single subtypes are calculated using the same cases and the same controls, which is checked by comparing 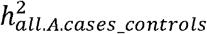 to 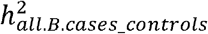 . GDIS uses several steps to compute this geometric representation of two subtype definitions:

#### Step 1: Estimate genetic correlations and compute angles between the subtypes

LDSC is used to compute *r* _*g*|*subA1*.*cases_con,subB1*.*cases_con*_, *r*_*g*|*subA2*.*cases_con,subB1*.*cases_con*_, *r* _*g*|*subA1*.*cases_con,subB2*.*cases_con*_, and *r* _*g*|*subA2*.*cases_con,subB2*.*cases_con*_, which are then converted to angles by taking the inverse cosine of these estimates.

#### Step 2: Rotate the triangle of the second subtype-definition

GDIS starts by overlaying both individual subtype-definition triangles on top of each other, such that the *all*.*cases_controls* line segments completely align. In principle, each of the four angles based on *r*_*g*|*subA1*.*cases_con,subBl*.*cases_con*_, *r*_*g*|*subA2*.*cases_con,subB1*.*cases_con*_, *r*_*g*|*subA1*.*cases_con,subB2*.*cases_con*_, and *r* _*g*|*subA2*.*cases_con,subB2*.*cases_con*_ can be used to rotate the second triangle; GDIS takes the angle corresponding to the genetic correlation estimate with the smallest standard error to perform the rotation. It will rotate the second triangle around the *all*.*cases_controls* line segment to match this angle (e.g., if *r*_*g*|*subA1*.*cases_controls,subB1*.*cases_controls*_ is used, the second triangle will be rotated in such a way that the angle between the line segments *subtypeA1*.*cases_controls* and *subtypeB1*.*cases_controls* corresponds to the angle from *r*_*g*|*subA1*.*cases_controls,subB1*.*cases_controls*_)

#### Supplementary validation analyses

If the *GWAS*_*subA1*.*cases_subA2*.*cases*_ and *GWAS*_*subB1*.*cases_subB2*.*cases*_ are available, GDIS will compare its derived value of *r*_*g*|*subA1*.*cases_subA2*.*cases,subB1*.*cases_subB2*.*cases*_ to the value estimated by LDSC. Specifically, it is determined what the difference between the GDIS-derived and the LDSC-estimated values are, and how the magnitude of this difference relates to the magnitude of the standard error of the LDSC-estimate value. We present these validation steps for our analyses in Figure 7 and Table S4.

#### Input filtering: check compatibility for geometrical representation

To check whether the computed values result in a geometrically realizable representation, GDIS first computes four helper triangles and determines whether they are geometrically valid (also see above). The helper triangles are formed by the points of the four different subtypes (subtypeA1-cases, subtypeA2-cases,subtypeB1-cases,subtypeB2-cases). Each helper triangle connects three subgroup points. The length of the lines of the helper triangles are computed in the same way as the lines of the helper triangle for the subtype with an external trait. When the sum of the two smallest lines are not longer than the longest line, the helper triangle is not valid. If one of the helper triangles is not valid, this indicates that the genetic correlations computed at step 1 are not compatible with each other and the two subtypes cannot be visualized together.

### Phenotype and genotype data

Data came from the UK Biobank (UKB)^37^, a population-based biobank containing genetic and phenotypic information on approximately 500,000 individuals. Mental health data were collected in multiple ways, including a mental health questionnaire that was available for a subset of individuals, linked hospital records and nurse interviews. The age of the individuals varies from 37 to 73 years and 54% of participants are female. Data were accessed under application 40310. Our main analyses are restricted to individuals of European ancestry for whom genetic and mental health questionnaire data^14^ were available (*N*=113,633).

### Phenotype definitions

#### MDD cases and controls

Our main analyses rely on previously defined cases and controls of lifetime MDD^14^. Diagnosis of MDD was based on the mental health questionnaire (MHQ), which included a modified version of the Composite International Diagnostic Interview Short Form (CIDI-SF). Individuals meeting criteria for a lifetime history of depression on the CIDI-SF were classified as MDD cases. Controls were those individuals that did not meet the lifetime history of depression based on the CIDI-SF, did not show signs of depression based on the Patient Health Questionnaire-9 (PHQ9), and did not indicate that they had ever been diagnosed with depression.

In secondary analyses (*N*=374,536), we used a broader definition of depression as used in the study by Nguyen et al^7^, which was based on either 1) meeting criteria for a lifetime or current diagnosis of MDD based on the CIDI-SF, a diagnostic instrument of MDD based on International Classification of Diseases version-10 from linked hospital records, or 2) having a probable depression (based on^38^), 3) self-report of a diagnosis of depression, or 4) exhibiting cardinal MD symptoms (prolonged loss of interest in normal activities or prolonged feelings of sadness or depression). Controls were semi-screened, i.e., individuals that had sought help for anxiety, nerves, tension, or depression or had used antidepressants, antipsychotics or lithium were excluded. Results of the secondary analyses using this broader MDD definition are reported in Figure S4. Genetic correlations between the GWASs based on the MHQ MDD definition and the broader definition of depression from^7^ can be found in Table S5.

#### Subtypes of MDD

We based our subtypes on previously proposed subtypes from^7^, with some slight adjustments to make the subtypes comparable to each other (i.e., in the original study, the different subtype-definitions were based on different definitions of cases and controls). We excluded cases that did not have data on all subtypes, to ensure that each subtype-definition was based on the same individuals (N subtype range 10,433-19,998; N controls: 83,202, Table S1). Genetic correlations of our subtypes based on the broader case-control definition of depression with the original GWASs from the study by Nguyen et al^7^ (also based on the broader case-control definition) can be found in Table S6.

- *MDD with or without childhood trauma:* In contrast to Nguyen et al^7^, we added the subtype of MDD with or without childhood trauma. MDD cases that had experienced at least one of the following five types of trauma were considered to have MDD with childhood trauma: physical abuse, emotional abuse, sexual abuse, emotional neglect, and/or physical neglect. Different cut-offs were used to determine the presence of each type of childhood trauma. Physical abuse was determined to be present if an individual indicated that this happened at least sometimes (UKB field ID: 20488). Emotional abuse was determined to be present if it happened at least sometimes (UKB field ID: 20487). Sexual abuse was determined to be present if it happened at least rarely (UKB field ID: 20490). Emotional neglect was determined to be present if an individual indicated to never, rarely or sometimes have felt loved as a child (UKB field ID: 20489). Physical neglect was determined to be present if an individual indicated they never, rarely or sometimes had someone to take them to a doctor if needed (UKB field ID: 20491).
- *MDD with or without comorbid anxiety*: Cases were considered to either have or not have comorbid anxiety based on self-reported anxiety (UKB field ID: 20544, 20002) or the presence of an ICD-diagnosis of anxiety (UKB field ID: 41202). In the original definition by Nguyen et al^7^, controls were excluded if they had an anxiety disorder. In this paper, the controls were not screened for anxiety, but remained the same across all subtypes (i.e., the MDD controls defined above).
- *MDD with or without early onset:* Individuals that reported having their first depressive episode before the age of 30 were considered to have MDD with an early onset, whereas individuals that have reported having their first depressive episode after the age of 30 were considered to have MDD without early onset (UKB field ID: 20433). In the original definition by Nguyen et al^7^ individuals with an onset before the age of 30 are considered to have MDD with an early onset and individuals with an onset after the age of 44 are considered individuals with a late onset of MDD. This definition was modified to a binary distinction, because the GDIS-visualization requires the combination of the subtypes to form the full set of all cases.
- *MDD with or without recurrent episodes:* Recurrence was based either on ICD codes (UKB field ID: 41202) specifying whether the depression was single episode or recurrent, on probable recurrent or single episode depression (UKB field ID: 20126) as defined by^38^, or on the self-reported lifetime number of depressed episodes (UKB field ID: 20442). If any of these three variables indicated that the person had more than one episode, they were considered to have the recurrent subtype. For the rest of the individuals, if any of the criteria indicated a single episode, they were considered to be of the single episode subtype.
- *MDD with or without high severity:* The original definition by Nguyen et al^7^ included the question on whether a case had mild or moderate recurrent depression (UKB field ID: 20126). However, because this question is also used in the single versus recurrent subtype, we changed the definition of this subtype. We based the definition on the number of CIDI symptoms that an individual with MDD endorsed (UKB field IDs: 20446-0.0, 20441-0.0, 20536-0.0, 20532-0.0, 20449-0.0, 20450-0.0, 20435-0.0, 20437-0.0). Individuals with 5 or 6 symptoms were defined as having MDD without high severity, while individuals with 7 or 8 symptoms were defined as having MDD with high severity. Note that only 8 questions are asked in the UKB and that the maximum number of symptoms is therefore 8 instead of all 9 CIDI symptoms.
- *MDD with or without sleep or weight increase:* Individuals that experienced weight increase or increased sleep during their worst depressive episode (UKB field IDs: 20536, 20534) were considered to have MDD with sleep or weight increase. This definition differs from the definition by Nguyen et al^7^ where the subtype is called MDD with atypical-like features, as they required individuals to have both increased weight and increased sleep. We made this definition less strict to increase the number of individuals with the increased sleep or weight subtype. Individuals that are considered to not have sleep or weight increase could have either experienced decreased sleep and/or weight or remained stable. Further subdividing this subtype would be of interest, but we lacked the sample size to do so.
- *MDD with or without suicidality:* Individuals with suicidality were those that indicated having thoughts of death during their worst depression (UKB field ID: 20437), or having recent thoughts of suicide or self-harm (either several days, more than half of the days or nearly every day; UKB field ID: 20513). In contrast to Nguyen et al^7^ we did not add the subtype of MDD with or without impairment, as a requirement for having MDD using the MHQ definition is that individuals need to experience impairment. We furthermore did not include the subtype of post-partum depression, as the full set of individuals with MDD cannot be as clearly separated into two subtypes (i.e., men and women that have not given birth yet cannot be placed into one of the subgroups) and the controls would need to be limited to women that have given birth as well, making the comparisons between subtypes problematic.

#### External traits

We investigated the genetic relation of the depression subtypes to 15 external traits (Table S2), including four dichotomous traits: anxiety disorder (ANX)^13^, bipolar disorder (BIP)^15^, major depressive disorder (MDD)^2^, schizophrenia (SCZ)^16^; four continuous traits: childhood trauma (CT)^17^, body mass index (BMI)^18^, educational attainment (EA)^19^, Wellbeing^20^, Neuroticism^21^; and six Genomic-SEM cross-disorder factors: substance use dependence factor^22^, compulsive factor^22^, internalizing factor^22^, neurodevelopmental factor^22^, thought disorders factor^22^, and a hierarchical factor representing overall liability to a psychiatric disorder^22^. GWAS sample sizes and heritabilities are reported in Table S2.

### Genome-wide association study

For each subtype-definition, we ran a GWAS of MDD cases with the feature vs. controls, MDD cases without the feature vs. controls and MDD cases with the feature vs. MDD cases without the feature.

We ran GWASs on related individuals using fastGWA, a mixed linear model for binary traits as implemented in GCTA v1.94.1^36^. We calculated the *N*_*effective*_ as follows: 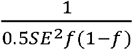, where is the allele frequency of the genetic variant and SE is the standard error of the effect of the variant^35^.

We extracted HapMap3 SNPs from genotypes imputed by the UKB (UKB field ID: 22828) and performed the following quality control: minor allele frequency (MAF) > .01, Hardy-Weinberg Equilibrium (HWE) p-value > 10^-10^ and missingness > 0.5. The genetic relationship between individuals was accounted for using a sparse genetic relationship matrix constructed with GCTA (see ref.^39^ for details). Covariates were age, sex, genotyping array and the first 10 genetic principal components (as determined in^39^).

As a sensitivity analysis to assess the impact of relatedness, we ran GWAS on unrelated individuals using PLINK 2^39^. We calculated the *N*_*effective*_ as follows: 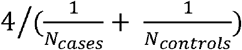 (refs.^33, 34^). We again used the HapMap3 imputed SNPs. Covariates were age, sex, genotyping array and the first 10 principal components. The genetic correlation results between the GWASs of related individuals with GWASs of unrelated individuals ranged from 0.977 to 1.362 (Table S7).

## Data Availability

All scripts used in the present study are available online at https://github.com/ABThijssen/GDIS/ and https://github.com/ABThijssen/Depression_heterogeneity/

## ACKNOWLEDGEMENTS

We thank Alkes Price for helpful and insightful discussions. A.B.T. is supported by a PhD scholarship from Amsterdam University Medical Center. A.B.T., W.J.P., M.B. and B.W.J.H.P. are supported by Stress in Action. The research project ‘Stress in Action’: www.stress-in-action.nl is financially supported by the Dutch Research Council and the Dutch Ministry of Education, Culture and Science (NWO gravitation grant number 024.005.010). The work of W.J.P. was partly financially supported by the Amsterdam Cohort Hub, which is part of the Sector Plan ‘Accelerating Health’ of the Dutch Ministry of Education, Culture and Science. UK Biobank: This study has been conducted using UK Biobank resource under application number 40310. Y.M. is supported by Amsterdam UMC (Starter Grant Ronde 2), Amsterdam Neuroscience (PoC funding 2024-2026), and the ImmunoMIND consortium, funded by UK Research and Innovation as part of the UK national Mental Health Platform. KJHV is supported by the Foundation Volksbond Rotterdam. M.B. is supported by an NWO VICI grant (VI.C.211.054).

## CONFLICT OF INTEREST STATEMENT

The authors report no conflicts of interest.

### URLs

UK Biobank: https://www.ukbiobank.ac.uk/

GDIS shiny app: https://GDIS.shinyapps.io/GDIS/

Package availability: https://github.com/ABThijssen/GDIS/

Code availability: https://github.com/ABThijssen/Depression_heterogeneity/

