## Supplementary Figures for "Estimating the genetic distance between subtypes of Major Depressive Disorder and their relationships with other traits using GDIS"

**Table of contents**
Figure S1. Extended figure on transforming heritabilities to distances and genetic correlations to angles.
Figure S2. Conversion of heritability on the liability scale to heritability on the observed scale.
Figure S3. GDIS visualizations of the seven major depression disorder subtypes for unrelated individuals.
Figure S4. GDIS visualizations of the seven major depression disorder subtypes with different case-control definition.
Figure S5. Side views of the GDIS visualizations of the relations between two subtype-definitions.
Figure S6. Genetic and phenotypic correlations between the subtypes.

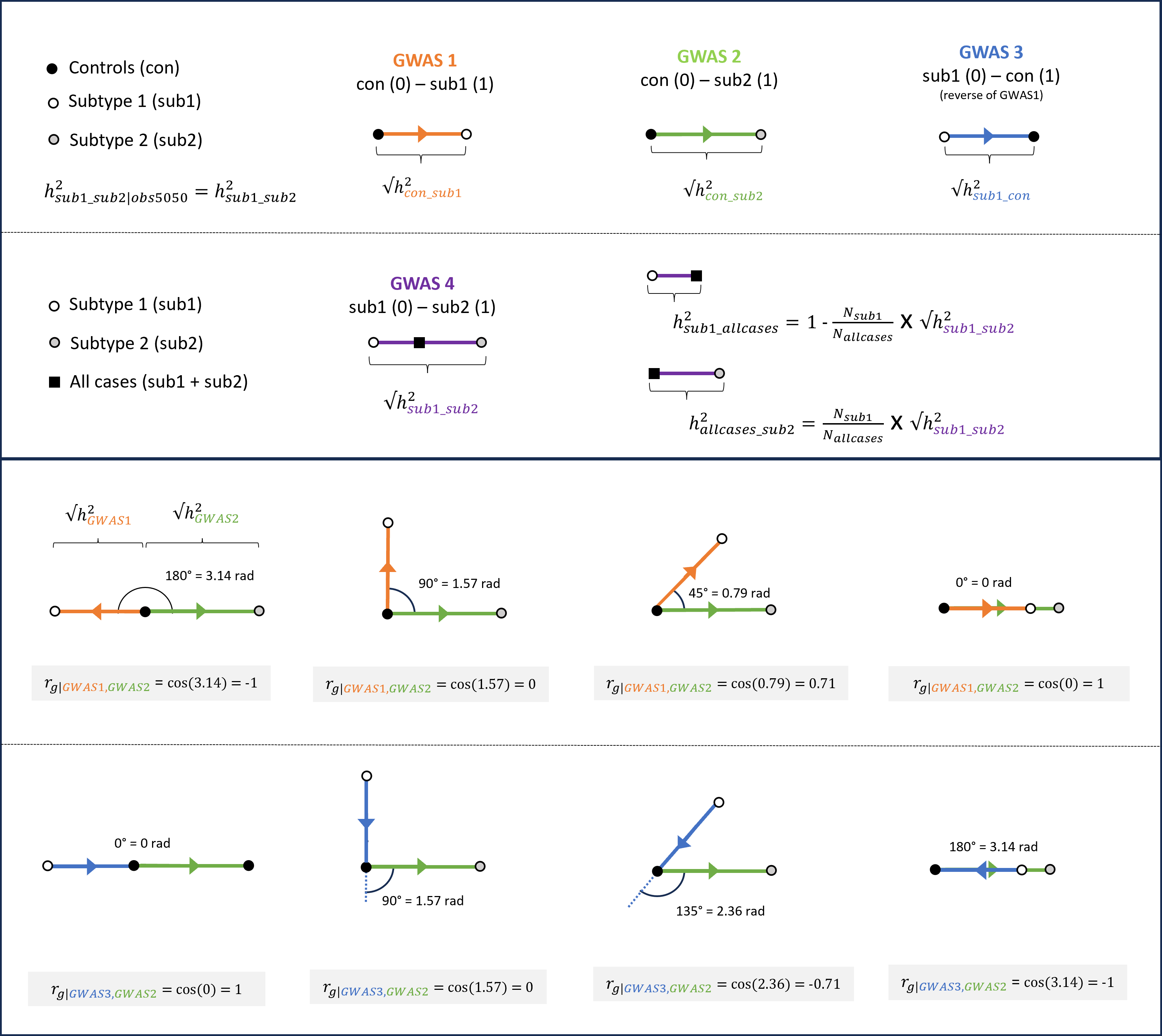
 **Figure S1. Extended figure on transforming heritabilities to distances and genetic correlations to angles.** The square root of the heritability on the observed scale with 50/50 ascertainment ($h_{obs|50/50}^{2}$) provides a Euclidian distance measure between the two compared groups; the line segment follows the direction from the GWAS of the class coded as 0 to the class coded as 1 (the direction is relevant for the angle). The group all-cases lies on the line segment between subtype1-cases and subtype2-cases, where the distance between all-cases and subtype2-cases is inversely proportional to the proportion of subtype2-cases among all-cases. The genetic correlation between two GWASs corresponds to the inverse cosine of the angle formed by the line segments of these comparisons. Depending on the direction of the comparison, the angle of interest is either the smaller angle formed directly between the lines, or the larger complementary angle. Note that the inverse cosine gives the angle in radians, but we report the angle in degrees.

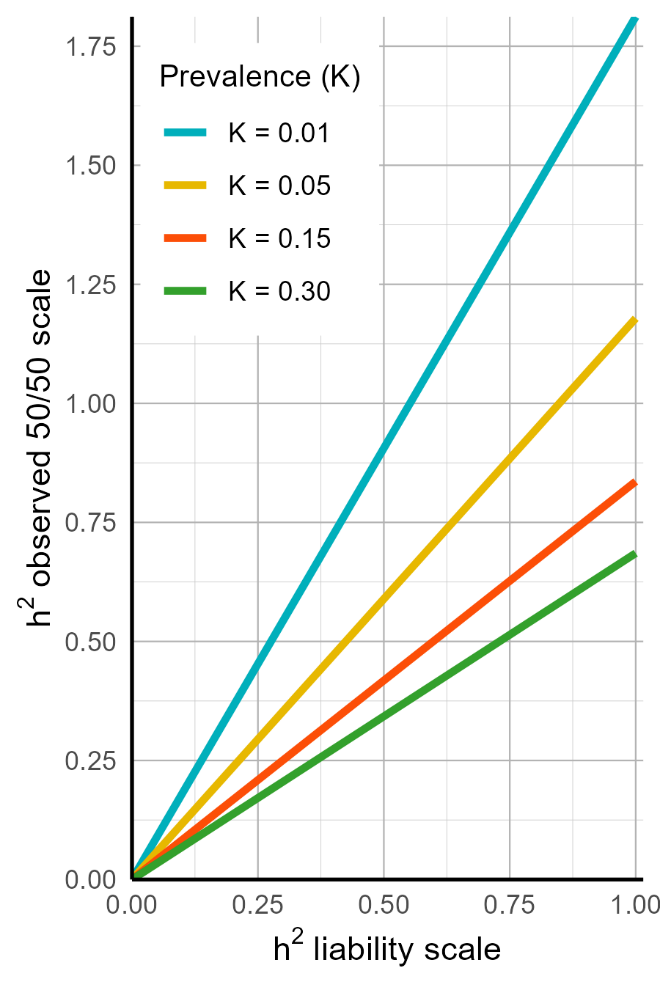
 **Figure S2. Conversion of heritability on the liability scale to heritability on the observed scale.** Examples of conversion of heritability on the liability scale to heritability on the observed scale are given for the population prevalences *K* = 0.01, *K* = 0.05, *K* = 0.15, and *K* = 0.30. The conversion function from (ref Lee et al., 2011) is $h_{observed}^{2}= \frac{h_{liability}^{2}*P*\left( 1-P \right)* {zv}^{2}}{K^{2}*{(1-K)}^{2}}$ , where *P* is the sample prevalence (which is 0.5 for heritabilities on the scale of 50:50 case-control ascertainment), *K* is the population prevalence and *zv* is the height of the standard normal curve at the liability threshold corresponding to prevalence.

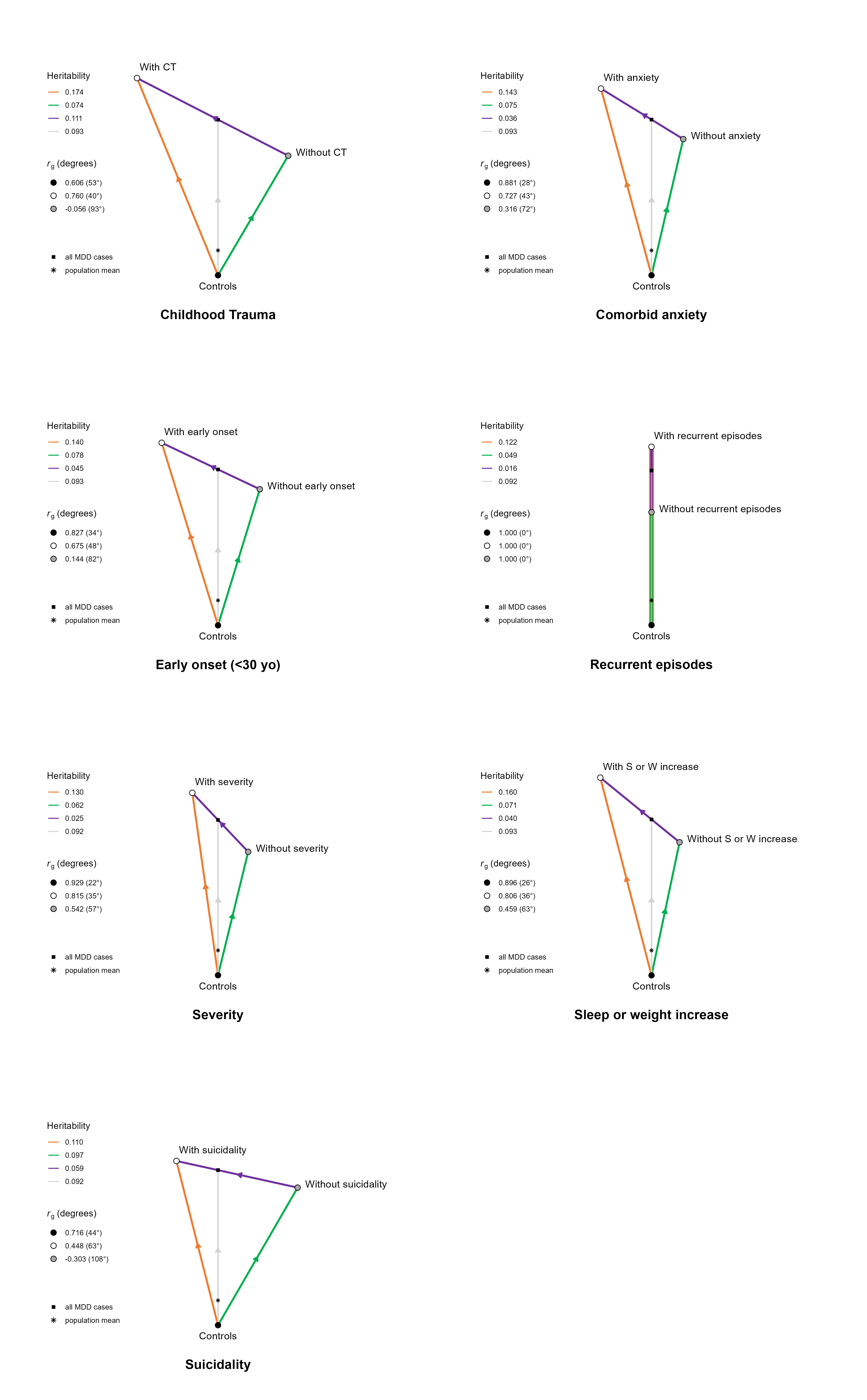
 **Figure S3. GDIS visualizations of the seven major depression disorder subtypes for unrelated individuals.** We present the GDIS visualization of the following seven subtypes of Major Depressive Disorder (MDD): with (subtype1-cases)/ without (subtype2-cases) childhood trauma, comorbid anxiety, early onset (< 30 yo), recurrent episodes, severity, sleep or weight increase, suicidality. These secondary analyses show similar findings to those presented in Figure 3.

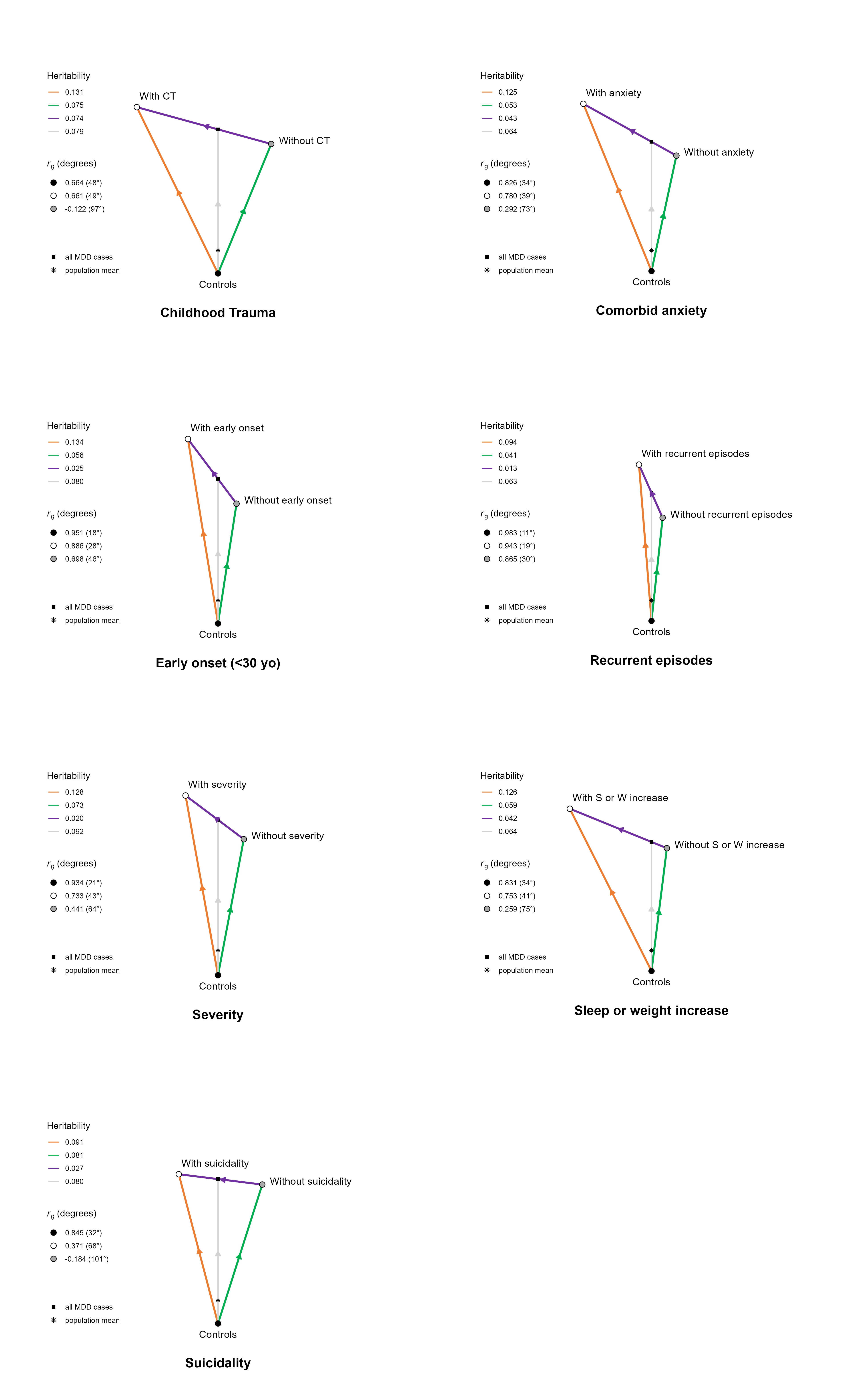
 **Figure S4. GDIS visualizations of the seven major depression disorder subtypes with different case-control definition.** We present the GDIS visualization of the following seven subtypes of Major Depressive Disorder (MDD): with (subtype1-cases)/ without (subtype2-cases) childhood trauma, comorbid anxiety, early onset (< 30 yo), recurrent episodes, severity, sleep or weight increase, suicidality. The case definition was based on a broad depression definition from the Nguyen et al. study. These secondary analyses show that findings are overall comparable to those from Figure 3, with slightly more pronounced differences between the visualizations of with/without early onset and with/without sleep or weight increase.

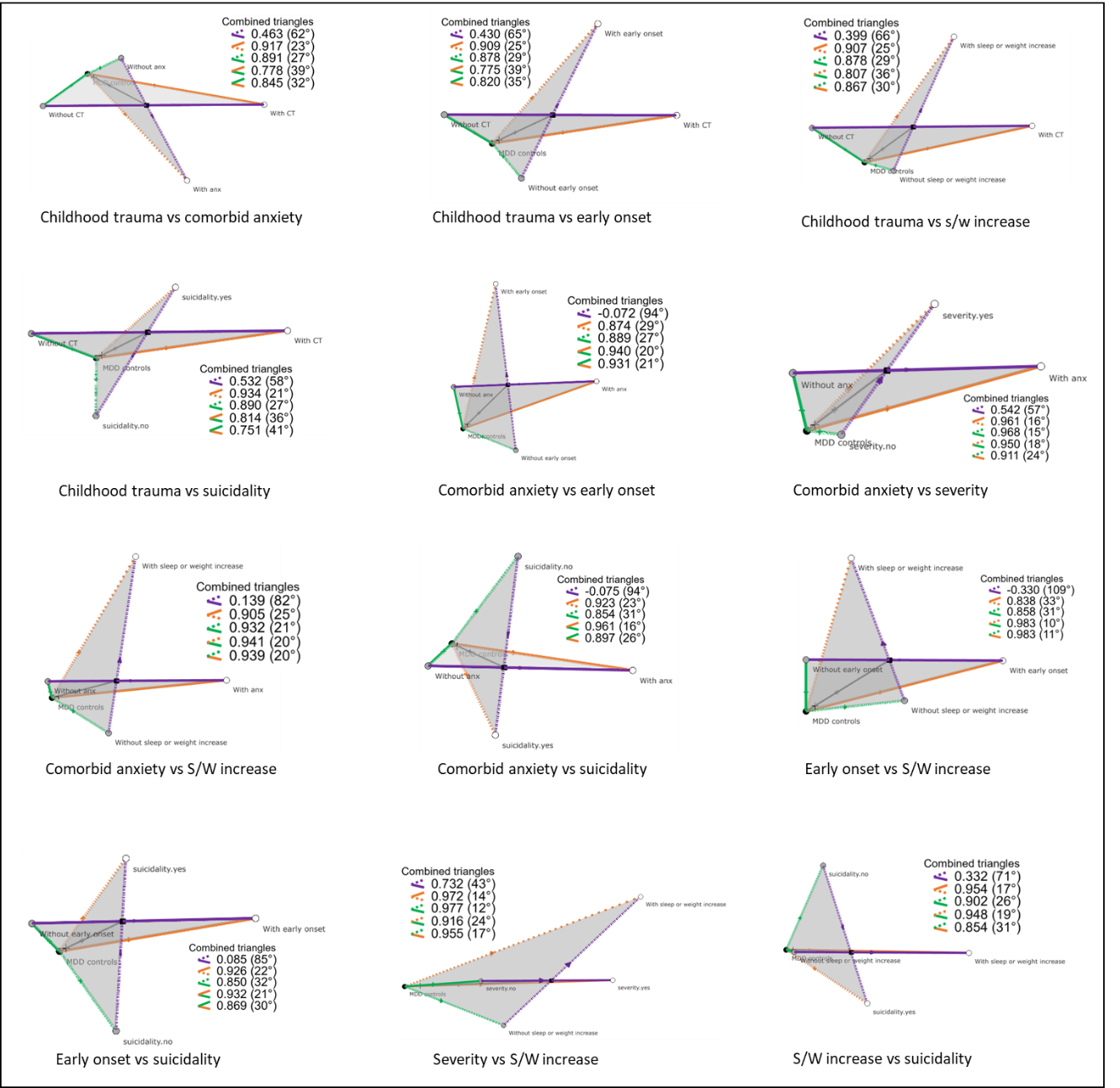
 **Figure S5. Side views of the GDIS visualizations of the relations between two subtype-definitions.** 2D views of the 3D visualizations of the relations between two subtype-definitions. The population mean and all major depressive disorder (MDD) cases are denoted by the cross and the black cube, respectively. The genetic correlation between two genome-wide association studies (GWASs) are denoted in the legend by the angle with the two colors representing the GWASs. All subtype-definitions were compared to each other (omitting all of the combinations with recurrence and most of the combinations with severity, as these were not geometrically valid. S/W increase = Sleep or weight increase. 3D visualizations can be interactively accessed at: https://gdis.shinyapps.io/gdis/.

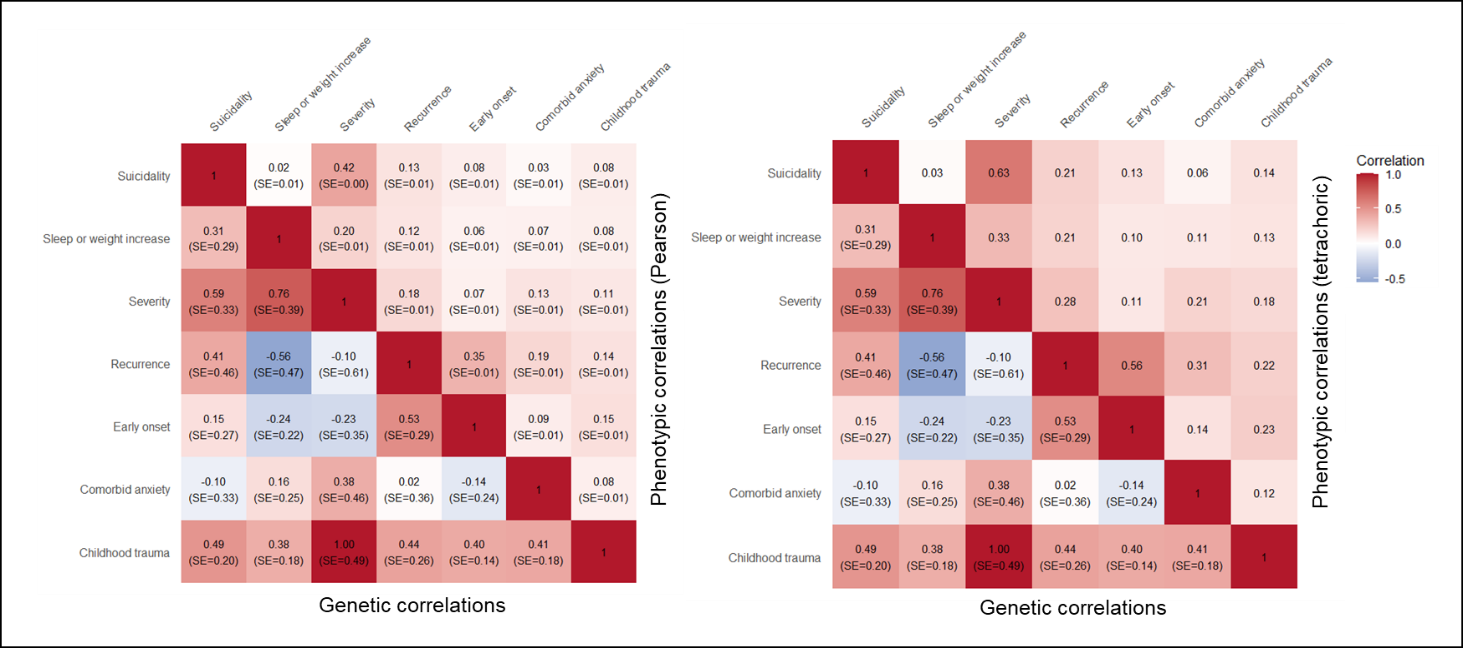
 **Figure S6. Genetic and phenotypic correlations between the subtypes.** Genetic correlations were computed using LDSC on ${GWAS}_{sub1.cases\_sub2.cases}$ for each subtype-definition combination. Phenotypic correlations are on the same subset of individuals and are computed using Pearson (left) and tetrachoric (right) correlations. Pearson and Spearman correlations give the same result due to the binary nature of the subtypes. Tetrachoric correlations do not come with standard errors.
